# Determining the optimal COVID-19 policy response using agent-based modelling linked to health and cost modelling: Case study for Victoria, Australia

**DOI:** 10.1101/2021.01.11.21249630

**Authors:** Tony Blakely, Jason Thompson, Laxman Bablani, Patrick Andersen, Driss Ait Ouakrim, Natalie Carvalho, Patrick Abraham, Marie-Anne Boujaoude, Ameera Katar, Edifofon Akpan, Nick Wilson, Mark Stevenson

## Abstract

**Importance:** Determining the best policy on social restrictions and lockdowns for the COVID-19 pandemic is challenging.

**Objective:** To determine the optimal policy response ranging from aggressive and moderate elimination, tight suppression (aiming for 1 to 5 cases per million per day) and loose suppression (5 to 25 cases per million per day).

**Design:** Two simulation models in series: an agent-based model to estimate daily SARS-CoV-2 infection rates and time in four stages of social restrictions; a proportional multistate lifetable model to estimate long-run health impacts (health adjusted life years (HALYs) arising from SARS-CoV-2) and costs (health systems, and health system plus GDP).

The net monetary benefit (NMB) of each policy option at varying willingness to pay (WTP) per HALY was calculated: NMB = HALYs × WTP – cost. The optimal policy response was that with the highest NMB.

**Setting and participants:** The State of Victoria, Australia, using simulation modeling of all residents.

**Main Outcome and Measures:** SARS-CoV-2 infection rates, time under various stages of restrictions, HALYs, health expenditure and GDP losses.

**Results:** Aggressive elimination resulted in the highest percentage of days with the lowest level of restrictions (median 31.7%, 90% simulation interval 6.6% to 64.4%). However, days in hard lockdown were similar across all four strategies (medians 27.5% to 36.1%).

HALY losses (compared to a no-COVID-19 scenario) were similar for aggressive elimination (286, 219 to 389) and moderate elimination (314, 228 to 413), and nearly eight and 40-times higher for tight and loose suppression. The median GDP loss was least for moderate elimination ($US41.7 billion, $29.0 to $63.6 billion), but there was substantial overlap in simulation intervals between the four strategies.

From a health system perspective aggressive elimination was optimal in 64% of simulations above a willingness to pay of $15,000 per HALY, followed by moderate elimination in 35% of simulations.

Moderate elimination was optimal from a partial societal perspective in half the simulations followed by aggressive elimination in a quarter.

Shortening the pandemic duration to 6 months saw loose suppression become preferable under a partial societal perspective.

**Conclusions and Relevance:** Elimination strategies were preferable over a 1-year pandemic duration.

**Funding:** Anonymous philanthropic donation to the University of Melbourne.

**Key points:** *Question:* To determine the optimal of four policy responses to COVID-19 in the State of Victoria, Australia (aggressive and moderate elimination, tight suppression (aiming for 1 to 5 cases per million per day) and loose suppression (5 to 25 cases per million per day), based on estimated future health loss and costs from both a health system and partial societal perspective.

*Findings:* From a health system perspective aggressive elimination was optimal in 64% of simulations above a willingness to pay of $15,000 per HALY, followed by moderate elimination in 35% of simulations. Moderate elimination was optimal from a partial societal perspective (i.e., including GDP losses) in half the simulations followed by aggressive elimination in a quarter.

*Meaning:* Whilst there is considerable uncertainty in outcomes for all the four policy options, the two elimination options are usually optimal from both a health system and a partial societal (health expenditure plus GDP cost) perspective.

## Introduction

There is no ‘right’ COVID-19 approach for all countries to follow. Rather, each country will devise an approach given its infection load, its ability to manage borders and quarantine, vulnerability and age structure of its population, health system capacity, economic and other resources, social values and preferences^1^ – and more recently timelines and progress to vaccination. One way to support this challenging decision-making is to use an integrated assessment of health and economic outcomes. Such an optimisation approach is often implicit in commentaries, with phrases such as “ensuring the cure is not worse than the disease”. But seldom is this balance or optimization explicitly defined and empirically addressed.

To this point in the pandemic, a standard ‘cost-effectiveness’ analysis resting on epidemiological modelling has not been widely used with the exception of cost-effectiveness and cost-benefit studies looking at a narrow range of policy measures or focused on a particular population group (e.g., ^2,3^ and reviews ^4,5^). Undertaking cost effectiveness studies in a pandemic is challenging for reasons such as which perspective to use (health system only, or societal) and uncertainty in many inputs – yet be it implicit or explicit, cost-effectiveness does feature in decision-making.^5^

In this paper we apply an integrated epidemiological and economic modelling approach to estimate the optimal COVID-19 pandemic control strategy for varying willingness-to-pay (WTP) per health adjusted life year (HALY) thresholds in a single high-income jurisdiction – the state of Victoria, Australia, as it was coming out of its second wave from 1 September. The four policy options include continuing stringent social and economic controls in an effort to achieve elimination of local transmission (‘aggressive elimination’ and ‘moderate elimination’ strategies), compared to ‘living with the virus’: ‘tight suppression’ as in some East Asian countries with low daily rates that the public health workforce can mostly manage with contract tracing; and ‘loose suppression’ with daily rates within health services capacity (e.g. countries like Spain, the UK and France between their first and second waves). We use both a health system perspective and a partial societal perspective (the latter adding GDP losses to health expenditure).

## Methods

### Conceptualisation and specification of policy scenarios

Most countries have some form of stages, ‘tiers’, or levels of control measures that a country or jurisdiction escalates up or de-escalates down depending on SARS-CoV-2 infection rates. Supplementary Table 1 shows the stages as applied in this study. Stage 1 has minimal restrictions, and by stage 4 there are marked restrictions on who are essential workers and what is considered an essential workplace with stay-at-home order applying to remaining citizens (i.e., stage 4 equates to ‘hard lockdown’). The rules encoded in the ABM to escalate and de-escalate stages are shown in Supplementary Table 1 – and for ‘tight suppression’ (as one example) in Supplementary Figure 1. We assume that mandatory mass-mask-wearing will be retained in stages 2 to 4, and in stage 1a and 1b will remain mandatory in public transport and busy indoor settings where physical distancing is difficult.

### Agent-based model

Further details on the ABM specification, validation and calibration are given elsewhere.^6,7^ Briefly, the model is a generic COVID-19 model of 2,500 agents that is then scaled up to the population of interest, here the state of Victoria with a population of 6.4 million. The ABM has a daily cycle length. Each agent ‘moves’ in a two-dimensional space, contacting other agents, with opportunities for between-agent infection given transmission rates and contact duration. The model has been calibrated to the first waves in NZ and Australia, has performed well in prediction (noting inherent stochastic uncertainty) ^6^, and has been used by the Victorian Government to help develop its RoadMap out of the second wave in Victoria.^8,9^ Supplementary Table 3 and 4 give the parameterisation of stages in the ABM.

### GDP and unemployment impacts

Each stage was associated with a GDP impact per week compared to business-as-usual or no COVID-19 (Stage 1 US$0.535 billion; Stage 1b US$0.6 billion; Stage 2 US$0.725 billion; Stage 3 US$1.275 billion; Stage 4 US$2.61 billion; Supplementary Table 7 and Appendix 1 for details).

### Proportional multistate lifetable model

A PMSLT ^10,11^ consists of parallel cohort lifetables for every sex by five-year old age cohort in 2020, simulated for all-cause mortality and morbidity over the remainder of their lives. Beneath this main all-cause lifetable sit parallel disease lifetables that proportions of the cohort reside in based upon disease incidence, case fatality and remission rates. In this paper, we use a PMSLT adapted for COVID-19 with disease states for SARS-CoV-2 infection and (for sensitivity analyses only – below) three indirect consequences of COVID-19 policy responses through changes in GDP and unemployment rates – road traffic crash (RTC), depression and anxiety. The inputs to the PMSLT are given in Supplementary Table 5.

The PMSLT was run 100 times for each policy response scenario using a 30-day cycle length, with the monthly number of SARS-CoV-2 cases from each of the ABM simulation inputted. These infections were distributed by age to match the actual proportionate distribution by age of cases in Victoria (skewed to older ages due to outbreaks in residential aged care). The outputs of the PMSLT include incremental HALYs and net health expenditure due to SARS-CoV-2 cases and average citizen health expenditure among the living, incremental to the no-COVID-19 business-as-usual. Up-front health expenditure (e.g., ICU capacity, surveillance systems) was assumed to be a ‘sunk cost’ the same between scenarios. 90% uncertainty intervals about these two outputs capture both stochastic uncertainty (from the varying infection rates in the ABM) and input parameter uncertainty (in the PMSLT about variables such as hospitalisation rates from SARS-CoV-2, and to a lesser extent input parameter uncertainty in the ABM).

We then estimate the monetary benefit (NMB) approach for each of the 100 runs:

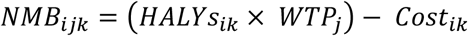

Where:

‐ i indexes the 100 iterations
‐ j indexes the WTP
‐ k indexes the four policy scenarios
‐ and Cost is the net health expenditure for the health system perspective analyses, and from the societal perspective adds GDP costs to health system costs.

Within each iteration i and WTP j, the policy scenario with the highest NMB is selected. Across all 100 iterations, each policy response k will have a probability of having the highest NMB, and the policy option with the highest probability is deemed ‘optimal’ at that WTP. Finally, these outputs can be shown as cost effectiveness acceptability curves.

### Sensitivity analyses

For HALYs and net health expenditure, they were re-estimated for 0% and 6% discount rates. Our base assumption of an intervention duration (i.e., the time to a vaccine) was 12 months; we reran the PMSLT for 6- and 18-month duration outputs from the ABM.

The main model only considered SARS-CoV-2 events. It seems likely that there are impacts of social restrictions on mental health (deleterious for depression and anxiety, but neutral for suicides) and reduced road traffic crash (RTC; beneficial). Ideally, one would estimate the counterfactual impacts on these health events at each level (stage) of COVID-19 policy response using published natural experiments; but this is a major task with, currently, inadequate data. Therefore, as a ‘simple’ sensitivity analysis we, first, for RTC used the association of mobility data with stages experienced in Victoria and in turn the association of mobility data with Victorian RTC rates (Appendix 4). Second, for depression and anxiety we assumed time spent in at stages 1a, 1b, 2, 3 and 4 had 2%, 4%, 6%, 8% and 10% increases in the age by sex prevalence of depression and anxiety, respectively.

Finally, a key rationale for tight suppression is that by keeping case numbers low, contact tracing is more effective, meaning one can function with lesser social and public health restrictions. We therefore reran the four policy scenarios incorporating a dynamic contract tracing variable that varied with the log of the number of cases per day (% of contacts identified per day = 0.586 – ln (cases per day) × 0.06213). Accordingly, the base case of 30% of contacts traced each day was assumed to apply at 100 cases per day, improving to 34% at 50 cases a day, 39% and 25 cases a day, 44% at 10 cases a day and 50% at 4 cases a day.

Findings are reported in accordance with the Consolidated Health Economic Evaluation Reporting Standards (CHEERS) checklist.^12^

## Results

### Impacts of each policy response strategy

The daily numbers of SARS-CoV-2 for 100 runs of each strategy are shown in Figure 1. All strategies saw a decrease in cases in the first two months, then marked stochastic variation in daily numbers (and hence varying times in stages 1a to 4 as shown in Figure 1 by varying the colour of each run’s trace).

**Figure 1.**
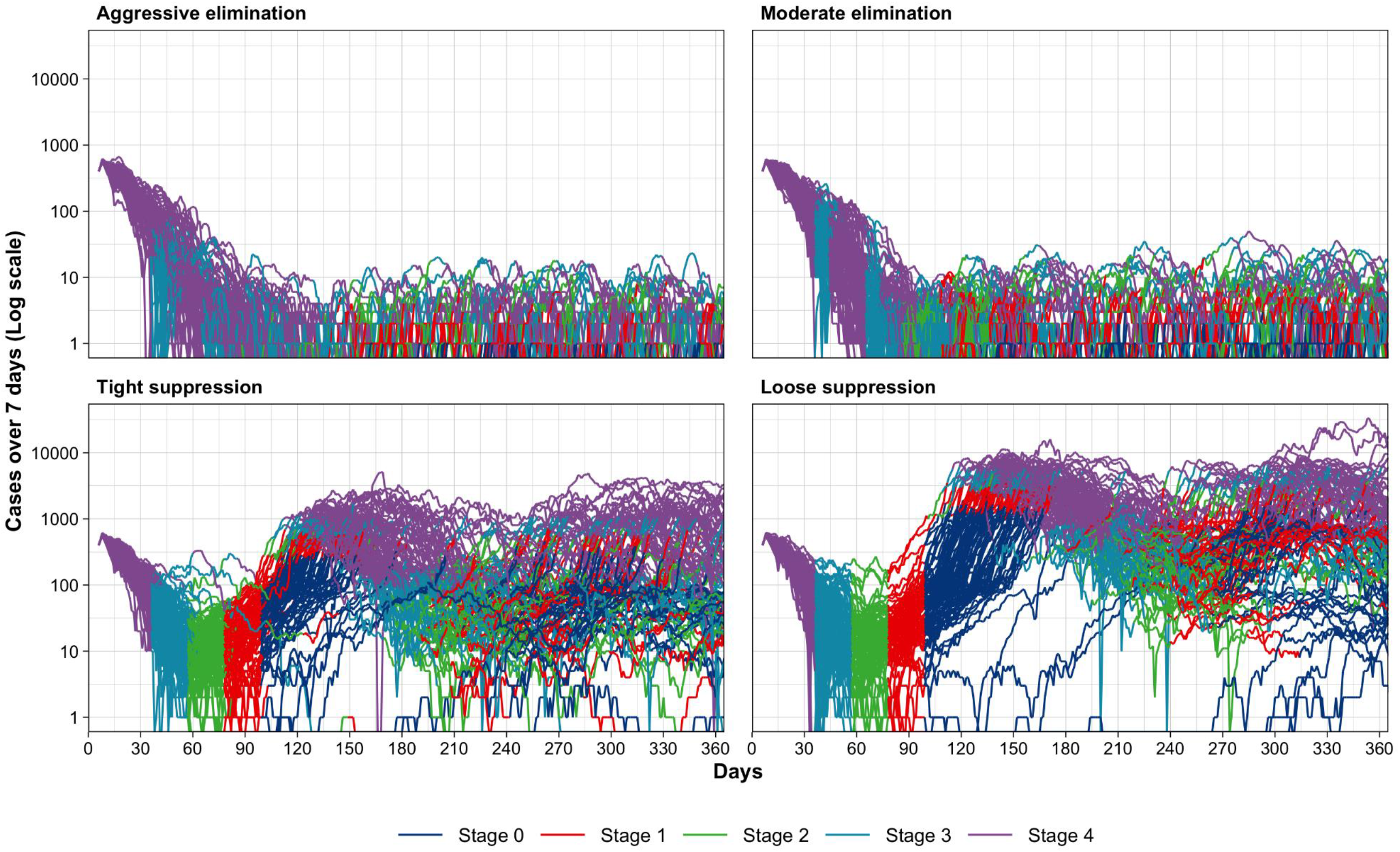
Daily cases per day for 100 runs of each of the four policy responses (colours of each line demonstrate the stage each run was in by day)

Table 1 shows the outputs from the ABM in terms of numbers of infections and numbers of days at varying policy stringency. The cumulative number of additional SARS-CoV-2 infections from the starting date over one year were lowest for aggressive elimination (median: 1,530, 90% simulation interval [SI]: 1,150 [i.e., 5th percentile] to 2,030 [i.e., 95th percentile, across the 100 runs]). Infections were highest at 55,900 for loose suppression (90% SI: 28,700 to 82,300).

**Table 1:** Outputs from ABM, and estimated GDP loss, for ‘best’ scenario: 12 months intervention or ABM time horizon (i.e., assumed vaccination available in 12 months); 1% probability per day of incursion of infected person into Victoria in elimination strategies.

|  | Strategy | a) Aggressive elimination | b) Moderate elimination | c) Tight Suppression | d) Loose suppression |
| --- | --- | --- | --- | --- | --- |
| SARS-CoV-2 cases (90% UI) | In first month | 1,330 (1,040 – 1,580) | 1,350 (1,110 – 1,600) | 1,260 (1,020 – 1,500) | 1,260 (1,090 – 1,540) |
|  | Month 2-6 incl | 173 (49 – 557) | 229 (50 – 642) | 4,570 (943 – 13,300) | 23,200 (119 – 37,700) |
|  | Month 7-12 incl | 18 (0 – 54) | 34 (0 – 104) | 5,640 (1,070 – 21,600) | 35,100 (6,160 – 48,900) |
|  | Month 13-18 incl | 13 (0 – 55) | 31 (0 – 93) | 4,160 (17 – 19,600) | 9,940 (128 – 71,800) |
|  | *Up to 12 months* | 1,530 (1,150 – 2,030) | 1,650 (1,240 – 2,110) | 11,700 (6,520 – 32,200) | 55,900 (28,700 – 82,300) |
|  | *Up to 18 months for scenario* | 1,540 (1,150 – 2,050) | 1,680 (1,280 – 2,180) | 17,900 (6,970 – 41,700) | 77,100 (35,900 – 129,000) |
| Percentage (90% UI) of days in each level | 1a | 21.7% (2.1% - 53.9%) | 31.7% (6.6% - 64.4%) | 15.1% (5.3% - 39.7%) | 16.3% (5.0% - 40.9%) |
|  | 1b | 16.3% (8.1% - 25.6%) | 11.9% (5.8% - 22.8%) | 13.9% (10.3% - 18.7%) | 14.4% (11.2% - 32.6%) |
|  | 2 | 12.1% (5.8% - 19.7%) | 13.8% (5.8% - 21.9%) | 12.8% (11.7% - 21.4%) | 11.7% (11.5% - 19.7%) |
|  | 3 | 18.3% (8.1% - 27.2%) | 16.4% (8.1% - 26.7%) | 15.7% (13.3% - 25.0%) | 14.7% (12.2% - 20.8%) |
|  | 4 | 30.3% (18.0% - 41.2%) | 27.5% (15.8% - 33.7%) | 34.3% (20.0% - 49.5%) | 36.1% (22.4% - 46.2%) |
| Estimated GDP loss (90% UI) | In first month | 7.9 (5.8 – 10.8) | 7.9 (5.8 – 10.8) | 7.9 (5.8 – 10.8) | 7.9 (5.8 – 10.8) |
|  | Month 2-6 incl | 22.6 (15.8 – 36.7) | 21.1 (14.0 – 33.6) | 19.3 (12.1 – 26.6) | 17.5 (11.1 – 25.2) |
|  | Month 7-12 incl | 15.6 (7.4 – 25.6) | 13.3 (7.0 – 22.8) | 23.2 (11.4 – 37.7) | 25.3 (11.6 – 34.8) |
|  | Month 13-18 incl | 15.4 (7.9 – 25.8) | 14.6 (7.7 – 22.5) | 21.3 (7.7 – 32.4) | 17.4 (7.4 – 33.1) |
|  | *Up to 12 months* | 46.5 (31.9 – 67.6) | 41.7 (29.0 – 63.6) | 50.9 (35.2 – 70.2) | 50.2 (35.1 – 72.5) |
|  | *Up to 18 months for scenario* | 61.3 (45.5 – 89.6) | 56.7 (39.9 – 79.2) | 71.7 (46.9 – 93.3) | 67.5 (44.3 – 97.4) |

Moderate elimination (31.7%, 6.6% to 64.4%) had the greatest percentage of days with the least restrictions (stage 1a) compared to 21.7% for aggressive suppression, 15% and 16% for tight and loose suppression. However, the percentage of days with maximum restrictions (stage 4) was similar across all strategies (medians ranging from 27.5% for moderate elimination to 36.1% for loose suppression – but with wide overlapping 90% SIs). Consequently, given much of the total loss of GDP is driven by Stage 4, whilst moderate elimination had the lowest median GDP loss up to 12 months of $US41.7 billion (90% SI 29.0 to 63.6), it was similar across strategies when allowing for the wide SIs.

Table 2 shows that deaths varied by stage pro-rata with the above infections. The crude case fatality rate is about 4% across scenarios, reflection the concentration of cases in aged care during the Victorian second wave. Lifetime HALY losses, discounted at 3% annually, were similar for aggressive elimination (286, 90% SI 219 to 389) and moderate elimination (314, 228 to 413), and 7 1/2 and 36-times higher for tight and loose suppression (Table 2). Net health expenditure differences compared to BAU varied over the time horizon, with expenditure in the first year increasing by $US2.71 million for aggressive elimination ($1.49 to $3.82 million) and up to $117 million for loose suppression ($50 to $214 million). However, in out-years health expenditure decreases compared to BAU in all strategies due to fewer people being alive incurring increasing health expenditure as they age.

**Table 2:** Estimate incremental health loss (HALYs) loss compared to BAU (i.e., no SARS-CoV-2 pandemic) and additional health expenditure (3% discount rate)

|  | Strategy | a) Aggressive elimination | b) Moderate elimination | c) Tight Suppression | d) Loose suppression |
| --- | --- | --- | --- | --- | --- |
| SARS-CoV-2 deaths | 1<sup>st</sup> yr. | 58 (46, 82) | 64 (46, 88) | 483 (245, 1,358) | 2,249 (1,022, 3,555) |
| HALYs loss (90% UI) | 1<sup>st</sup> yr. | 38 (29, 50) | 41 (30, 54) | 156 (98, 364) | 672 (337, 954) |
|  | 2<sup>nd</sup> yr. | 71 (54, 94) | 76 (56, 101) | 436 (246, 1,120) | 1,990 (1,020, 2,960) |
|  | Full 20 years | 272 (204, 363) | 289 (215, 386) | 1,950 (1,100, 6,100) | 11,900 (4,730, 15,500) |
|  | Rest of lifetime | 286 (219, 389) | 314 (228, 413) | 2,263 (1,180, 6,550) | 11,000 (5,030, 16,700) |
| Net health expenditure (in \$US millions; 90% UI) | 1<sup>st</sup> yr. | 2.71 (1.49, 3.82) | 2.88 (1.69, 4.61) | 24.6 (10.4, 75.3) | 117 (50, 214) |
|  | 2<sup>nd</sup> yr. | 2.00 (0.994, 2.99) | 2.10 (1.12, 3.61) | 19.2 (7.19, 59.1) | 89.0 (33.1, 169) |
|  | Full 20 years | -2.54 (-3.95, -1.68) | -2.77 (-3.88, -1.79) | -18.5 (-51.5, -8.23) | -90.2 (-153, -30.6) |
|  | Rest of lifetime | -2.99 (-4.45, -2.00) | -3.19 (-4.51, -2.22) | -22.4 (-60.1, -9.92) | -107 (-175, -41.7) |

### Differences between policy responses

Table 3 shows the difference <u>between</u> policy response options, for the median and percentiles of the differences within each of the 100 runs. Unsurprisingly, there were clear differences in health loss with no overlap in 90% SIs for the two elimination options compared to the two suppression options.

**Table 3:** Differences in main outputs between the policies for 12-month intervention period (90% UI of the differences calculated within each run, ensuring each within-run comparison is subject only to stochastic variability and PMSLT input parameter uncertainty.

| Strategy | a) Aggressive elimination | b) Moderate elimination | c) Tight Suppression | d) Loose suppression |
| --- | --- | --- | --- | --- |
| ***Compared to aggressive elimination*** |  |  |  |  |
| HALYs <u>loss</u> lifetime | 0 (ref) | 21 (-90 to 129) | 1927 (871 to 6249) | 10661 (4776 to 16396) |
| Net health expenditure lifetime (\$US millions) | 0 (ref) | -0.2 (-1.3 to 0.9) | -19.2 (-57.4 to -7.4) | -103.2 (-172.8 to -40.2) |
| Estimated GDP <u>loss</u> (\$US billions) | 0 (ref) | -5.2 (-18.6 to 11.0) | 3.3 (-12.1 to 23.6) | 4.3 (-15.9 to 19.4) |
| ***Probability of being largest across policies*** |  |  |  |  |
| HALYs <u>loss</u> lifetime | 0% | 0% | 7% | 93% |
| Net health expenditure lifetime | 62% | 37% | 0% | 1% |
| Estimated GDP <u>loss</u> (\$US billions) | 19% | 4% | 33% | 44% |
| ***Probability of being smallest across policies*** |  |  |  |  |
| HALYs <u>loss</u> lifetime | 63% | 36% | 0% | 1% |
| Net health expenditure lifetime | 0% | 0% | 7% | 93% |
| Estimated GDP <u>loss</u> | 23% | 49% | 14% | 14% |

There is considerable stochasticity and overlap between policies in GDP loss, due to the elimination strategies using stage 3 and 4 (where GDP loss compared to BAU is maximal) more readily and early, while the suppression strategies have to move into Stages 3 and 4 frequently to keep infection and case rates within the target range. The net result is widely overlapping SIs in GDP loss for the four strategies (top panel of Table 3). Therefore, we also determine the percentage of runs in which GDP loss was the greatest: loose suppression had the greatest GDP loss in 44% of simulations, followed by tight suppression at 33% and aggressive elimination at 19% (middle panel of Table 3). Whilst not an exact mirror image, the percentage of runs in which GDP loss was the least was 49% for moderate elimination, followed by aggressive elimination at 23% and both suppression options at 14% each (bottom panel of Table 3).

Figure 2 shows the cost effectiveness acceptability curves for the four strategies. Interventions tend to be considered cost-effective up to approximately GDP per capita per HALY in high-income countries, which is about US$55,000 for Australia; for WTP greater than $15,000 per HALY, aggressive elimination is optimal in 64% of simulations, followed by moderate elimination in 35% of simulations.

**Figure 2.**
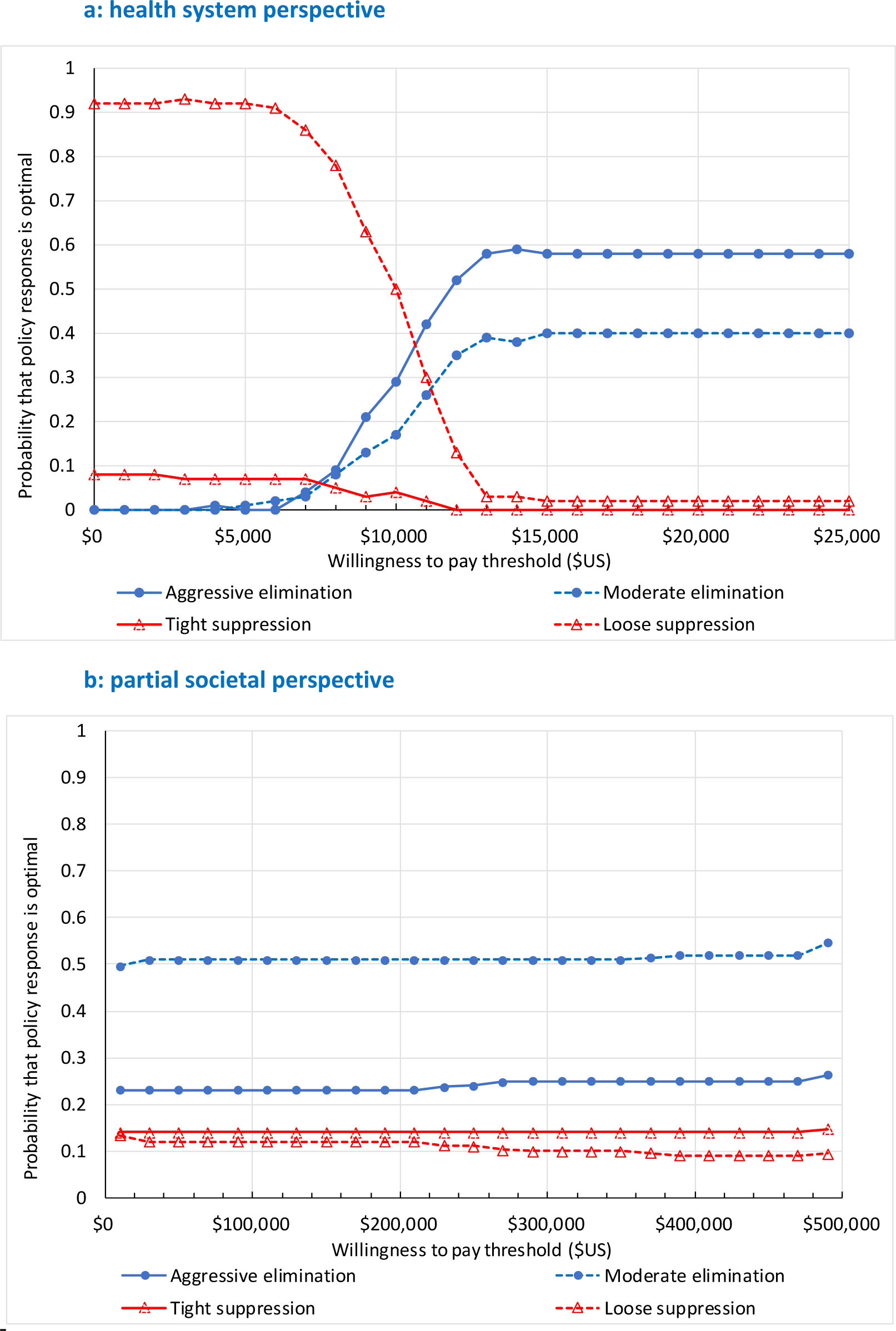
Cost effectiveness acceptability curves:

From a partial societal perspective (i.e., now treating costs as that in the health system plus GDP (which largely means GDP alone because it is orders of magnitude greater than the health expenditure impacts)), moderate elimination is optimal for half the simulations regardless of WTP, and aggressive elimination is optimal a quarter of the time. There is, however, a 10% to 15% probability of either tight or loose suppression being optimal, highlighting the uncertainty in estimates.

### Sensitivity analyses

Results for the sensitivity analyses are shown in Supplementary Table 6, all conducted (including the baseline) using the median number of infections from the ABM propagated through the PMSLT as one ‘expected or median value’ run. (Monte Carlo simulation was not feasible for sensitivity analyses due to long model run times.) Varying the discount rate to 0% and 6% altered HALYs and costs predictably and did not alter relativities between the policies. Also shown in Supplementary Table 6 is which policy is optimal (i.e., highest net monetary benefit) for the one median value run: from a health system perspective aggressive elimination is optimal above a WTP of $15,000 per HALY regardless of the discount rate, and moderate elimination is always optimal from a partial societal perspective.

Lessening the intervention duration to 6 months (i.e., as though the pandemic was over at six months due to early provision of vaccination), or lengthening it to 18 months duration, unsurprisingly decreased and increased the health losses and GDP losses, respectively. It also resulted in loose suppression displacing moderate elimination as optimal from a partial societal perspective for a 6-month intervention duration (Supplementary Table 6 and Supplementary Figure 2).

Including RTC in the model (in addition to just SARS-CoV-2 infection) saw health gains of about 900 HALYs for both elimination strategies. Conversely, including depression and anxiety saw further increases the health loss. Noting the speculative nature of these disease-scenario analyses, the combined effect of adding in RTC, depression and anxiety was health gains in the two elimination strategies (due to RTC decreases tipping the balance), but little relative impact on the health loss for loose suppression (due to SARS-CoV-2 health loss being much larger than any countervailing force of RTC). Nevertheless, the optimal strategy from a health system perspective remained one of the elimination strategies and from a partial societal perspective was always moderate elimination.

## Discussion

This study assesses both the health and economic impacts of COVID-19 policy response strategies. There are several important findings. First, there is large stochastic variation in SARS-CoV-2 infections between runs of the same strategy (Figure 1). Second, there was little difference in the median GDP loss across strategies – as elimination strategies use Stage 3 and 4 lockdowns at lower thresholds, whereas suppression strategies are forced to use Stage 3 and 4 to keep infections in the target range. Further, the GDP impacts are very uncertain. Comparing across countries, however, Baker et al found some suggestion that GDP loss during 2020 as estimated by the IMF has been lower in countries using elimination as opposed to suppression strategies.^13^ Loose suppression in our study had the largest probability (44%) of having the highest GDP loss, tight suppression had a 33% probability and aggressive elimination still had a 19% probability of the highest GDP loss. Third, the strategy with the highest net monetary benefit (and hence ‘optimal’ on cost-effectiveness grounds) strategy was aggressive elimination from a health system perspective and moderate elimination from a partial societal perspective; importantly, the two elimination strategies were the two most optimal strategies at usual WTP thresholds. Fourth, reducing the intervention duration from 12 to six months (approximating most people vaccinated in six months) saw loose suppression displace moderate elimination as optimal from a partial societal perspective – offering some support for countries that have high rates currently (e.g., UK, USA, Spain, France) to ‘ride it out’ with short lock-downs to keep infection rates in check until vaccination coverage is high.

Compared to the scant cost-effectiveness literature on COVID-19 policy responses so far ^4,5^, our study offers improvements. First, to the best of our knowledge our study is the only one that uses an ABM to simulate dynamic policy regimes that move up and down stages based on triggers of daily case numbers and captures the large stochastic variation in how case numbers evolve. Other model frameworks use rapid approaches ^14^, decision-tree and Markov or a variation of a dynamic (SIR-style) model which allow for only limited complexity (e.g., ^3^). Next, our consideration of both health costs and GDP impacts resulting from social policies put into place take us beyond a limited health sector perspective.^15^ A previous Australian analysis has also found that elimination is better for both health and economic outcomes ^16^ but was not as sophisticated as our study in allowing for dynamic policy settings, other disease sequalae, and viral reincursion (due to border breaches). Interestingly, our results echo somewhat a US study finding that social distancing is a cost-effective strategy relative to herd immunity if an effective therapy or vaccine can be introduced within a reasonable (under 12 months) timeframe.^14^

Our study also forces us to confront the reality that the outcomes of COVID-19 policy responses are very uncertain due to both stochastic uncertainty (i.e., chance occurrences of who contacts who, leading to highly variable epidemic curves) and uncertain knowledge about the impact of masks, apps, behaviour, and other interventions. Whilst stochastic uncertainty will persist despite improved knowledge, there are other aspects that may improve with better knowledge. First, contact tracing is improving as the pandemic progresses, which will favour elimination strategies assuming that contact tracing is most effective when caseloads are low. Second, it is strongly suspected (but hard to quantify) that unintended consequences of policy responses on non-SARS-CoV-2 health outcomes and long-COVID are important. For example, in plausible sensitivity analyses we used for RTC and depression and anxiety, the net health impact for elimination strategies with low SARS-CoV-2 caseloads can vary markedly in percentage terms.

Whilst the world is ramping up vaccine rollouts, this will take time and future resurgences are likely. Our study suggests that for those countries with an option to pursue or retain elimination an elimination strategy is optimal.

## Data Availability

The Victorian Department of Health
and Human Services provided Victorian SARS-CoV-2 notification data for calculation of hospitalisation,
ICU admission and fatality rates.

## Acknowledgements

We thank Professor Michael Baker for the contribution to defining stages, and Professor Rod McClure for assistance with ABM model building. The Victorian Department of Health and Human Services provided Victorian SARS-CoV-2 notification data for calculation of hospitalisation, ICU admission and fatality rates.

