## Supplementary Files for "Determining the optimal COVID-19 policy response using agent-based modelling linked to health and cost modelling: Case study for Victoria, Australia"

### Supplementary Tables and Figures

#### Supplementary Table 1: Conceptualisation and specification of the triggers to shift between stages by policy scenario.

| 1. Aggressive elimination † | 2. Moderate elimination † | 3. Tight suppression (aiming for 1 to 5 cases per million population per day)‡ | 4. Loose suppression (aiming for 5 to 25 cases per million population per day)‡ |
|---|---|---|---|
| **First, check for tightening (below rules applied on each day, regardless of Stage currently in)** | | | |
| If in Stage 3 or lower, and average >7.5 cases per day for last 7 days, go to Stage 4<br>Else:<br>If in Stage 2 or lower, and average > 1.5 cases per day for last 7 days, go to Stage 3<br>Else:<br>If in Stage 1b or 1, and two or more cases in last 14 days, go to Stage 2<br>Else:<br>If in Stage 1, and any single case, go to Stage 1b<br>Else:<br>Stay in same stage. | If in Stage 3 or lower, and average >30 cases per day for last 7 days, go to-Stage 4<br>Else:<br>If in Stage 2 or lower, and average > 6 cases per day for last 7 days, go to Stage 3<br>Else:<br>If in Stage 1b or 1, and five or more cases in last 14 days, go to Stage 2<br>Else:<br>If in Stage 1, and two or more cases in last 14 days, go to Stage 1b<br>Else:<br>Stay in same stage | If >20 per million in last 7 days, go to Stage 4.<br>Else:<br>If > 10 per million in last 7 days, go to Stage 3.<br>Else:<br>If average of >5 per million in last 7 days, and > 6 days since last tightening, tighten 1 stage<br>Else:<br>Stay in same stage | If >100 per million per day in last 7 days, go to Stage 4.<br>Else:<br>If > 50 per million per day in last 7 days, go to Stage 3.<br>Else:<br>If average of >25 per million per day in last 7 days, and > 6 days since last tightening, tighten 1 stage<br>Else:<br>Stay in same stage |
| **Second, check for loosening** | | | |
| If in Stage 4, and average <5 cases per day for last 7 days, and > 20 days since last loosening, go to Stage 3<br>Else:<br>If in Stage 3, and average < 1 case per day for last 7 days, and > 20 days since last loosening, go to Stage 2<br>Else:<br>If in Stage 2, and zero cases for last 7 days, and > 20 days since last loosening, go to Stage 1b<br>Else:<br>If in Stage 1b, and zero cases for last 28 days and > 20 days since last loosening, go to Stage 1<br>Else:<br>Stay in same stage | If in Stage 4, and average <20 cases per day for last 7 days, and > 20 days since last loosening, go to Stage 3<br>Else:<br>If in Stage 3, and average < 5 cases per day for last 7 days, and > 20 days since last loosening, go to Stage 2<br>Else:<br>If in Stage 2, and <1 case per day for last 7 days, and > 20 days since last loosening, go to Stage 1b<br>Else:<br>If in Stage 1b, and zero cases for last 7 days and > 20 days since last loosening, go to Stage 1<br>Else:<br>Stay in same stage | If average of <2.5 per million in last 7 days, and > 20 days since last loosening, loosen 1 stage<br>Else:<br>Stay in same stage | If average of <12.5 per million per day in last 7 days, and > 20 days since last loosening, loosen 1 stage<br>Else:<br>Stay in same stage |

‡ 1 case per million per day equates to an expected 6.4 cases per day in Victoria. A flow diagram of tight suppression is given in Supplementary Figure 1.

#### Supplementary Table 2: Physical distancing and other measures by 'stage' as used in Victoria, Australia, for control of COVID-19

| Domain | Condition | Stage 1 | Stage 1b | Stage 2 | Stage 3 | Stage 4 |
|---|---|---|---|---|---|---|
| Stay at home | Number of reasons to leave home† | - | - | 5 | 4 | 4 |
| | Limit on range of movement | - | - | - | 5 km | 5km |
| | Time away from home | - | - | - | 2 hours | 1 hour |
| | Limit on the number of times you can go out per day | - | - | - | - | 1 |
| | Curfew (8:00pm – 5:00am) | No | No | No | No | Yes |
| | Work from home | Return to work | If you can | If you can | If you can | Stay at home, unless defined essential worker |
| Home visitors (non-household members) | Maximum number (N) of visitors | 100 | 20 | 5 | 0 | 0 |
| Outdoor gatherings | Maximum N of persons (including for physical activity / exercise) | 100 | 20 | 10 | 5 | 2 |
| Industries, education, hospitality facilities (% closed unless otherwise stated) | Major construction sites | 0% | 0% | 0% | 0% | 75% |
| | Small scale construction, e.g., residential (max number of people on site) | - | - | - | - | 5 |
| | Meat industry | 0% | 0% | 0% | 0% | 33% |
| | Poultry industry | 0% | 0% | 0% | 0% | 20% |
| | Seafood industry | 0% | 0% | 0% | 0% | 33% |
| | Manufacturing | 0% | 0% | 0% | Only to supply essential services | Only to supply essential services |
| | Warehousing & distribution centres | 0% | 0% | 0% | 0% | 0% |
| | Technical and further education, & University studies | Opening gradually | Opening gradually | Opening gradually | Mostly remote learning | Only remote learning |
| | Schools | Open | Open | Open | Closed (except to vulnerable children and children of permitted workers | Closed (except to vulnerable children and children of permitted workers |
| | Childcare & pre-school care | Open | Open | Open | Open | Closed (except to vulnerable children and children of |

| Domain | Condition | Stage 1 | Stage 1b | Stage 2 | Stage 3 | Stage 4 |
|---|---|---|---|---|---|---|
| | | | | | | permitted workers |
| | Hardware stores | 0% | 0% | 0% | 0% | Closed – exception to tradespeople |
| | Department stores | 0% | 0% | 0% | 0% | 100% |
| | Hairdressers & barbershops | 0% | 0% | 0% | 0% | 100% |
| | Beauty parlours & massage therapy | 0% | 0% | 100% | 100% | 100% |
| | Real estate auction – max N | 100 | 20 | 15 | 0 | 0 |
| | Accommodation services – Closed | No | No | No | Yes | Yes |
| | Café & restaurants – $m^2$ per person | 4 | 4 | - | - | - |
| | Café & restaurant – max N | 100 | 20 | 0 | 0 | 0 |
| | Café & restaurant – closed | No | No | Yes | Yes | Yes |
| | Food courts – Closed | No | Yes | Yes | Yes | Yes |
| | Pubs, clubs, casinos & nightclubs | No | Yes | Yes | Yes | Yes |
| | Cinemas & entertainment services | 100 | 20 | 0 | 0 | 0 |
| Places of worship | Closed | No | No | No | Yes | Yes |
| | Maximum N allowed | 100 | 22 | 12 | 0 | 0 |
| | $M^2$ per person | - | 4 | 4 | - | - |
| | Weddings – maximum N allowed | 100 | 23 | 13 | 5 | 0 |
| | Indoor funerals – max N allowed | 100 | 52 | 22 | 12 | 12 |
| | Outdoor funerals – max N allowed | 100 | 52 | 32 | 12 | 12 |
| Face covering ‡ | | No | In public transport and indoor environment if not with household members | In public transport and indoor environment if not with household members | Mandatory out of home, unless doing vigorous physical activity | Mandatory out of home, unless doing vigorous physical activity |
| Sporting activities | Indoor sports – $m^2$ per person | - | 4 | - | - | - |
| | Indoor sporting centres – max N | 100 | 20 | 0 | 0 | 0 |
| | Gym – max N | 100 | 20 | 0 | 0 | 0 |
| | Play centres - % closed | 0% | 0% | 100% | 100% | 100% |
| | Playgrounds - % closed | 0% | 0% | 100% | 100% | 100% |
| | Recreation activities (fishing, golf, boating, | 0% | 0% | 0% | Allowed with one person | 100% |

| Domain | Condition | Stage 1 | Stage 1b | Stage 2 | Stage 3 | Stage 4 |
|---|---|---|---|---|---|---|
| | tennis, surfing, drive range shooting) - % closed | | | | | |
| Aged care restrictions | Max N visitors at one time | 2 | 2 | 2 | 0 | 0 |
| | Max N of visits per day per resident | 2 | 2 | 2 | 0 | 0 |
| | Max total duration of visits (in hours) | 2 | 2 | 2 | 0 | 0 |
| | Face masks required of visitors | If asked | If asked | If asked | Mandatory | Mandatory |
| | Workers working at multiple facilities | Allowed | Allowed | Allowed | Not allowed | Not allowed |
| | Facemask required of workers | No | Mandatory | Mandatory | Mandatory | Mandatory |

† At Stage 3 and 4, four reasons are: for essential work; for shopping (e.g., groceries); to give or receive care; for physical activity. At Stage 2, seeing friends and family added.

‡ This face covering usage is how it played out with escalating application of stages in Victoria. Going forward, it is likely that face coverings will be mandatory at all Stages – perhaps only on public transport and in busy indoor public spaces in stages 1, 1b and 2.

#### Supplementary Table 3: Parameter estimates and 'agent' characteristics most relevant to current paper used in the agent-based model (for full details see source code and ODD protocol in footnote to this table)

| Key Parameters | Parameter Estimates (Policies 1, 2, 3, 4) |
|---|---|
| **Physical distancing** (% of people limiting movement and maintaining a distance of 1.5m in public, normal distribution)[¥ 1] | m = 85%, sd = 3% |
| **Physical distancing - time** (% of time that people successfully maintain a distance of 1.5m, normal distribution)[¥1] | m = 85%, sd = 3% |
| **Mean incubation period** (days, log-normal)[2] | m = 5.1, sd = 1.5 |
| **Mean illness period** (days, log-normal)[3] | m = 20.8, sd = 2 |
| **Mean adherence with isolation of infected cases** (beta distribution) [¥] | m = 93.3%<br>(beta 28, 2; median = 94.3%, SD = 4.5%) † |
| **Number of days after initial infection that new cases are reported**[¥*] | 6* |
| **Date of case simulation initialisation** (Day 0) | September 1st, 2020 |
| **Asymptomatic cases** (% of cases, normal distribution) [3,4] | m = 33%, sd = 3% |
| **Infectiousness of asymptomatic cases vs symptomatic cases (per contact,** normal distribution**)** [5] | m = 33%, sd = 6% |
| **Reduction in transmission risk per contact for people wearing facemasks** (beta distribution) **¥¥¥** | 77%[6] (beta 24.3, 8.08)† |
| **Seeded cases** | An initial volume of 2400 active cases were seeded into the model on day 0. This was followed by 7 days of 80 cases per day. |
| **COVID-Safe Electronic App Uptake** (normal distribution) | m = 30% |

| Agent Characteristics | Definition |
|---|---|
| **Infection status** | Susceptible, Infected, recovered, deceased |
| **Time now** | The number of days (integer) since an infected person first became infected with SARS-CoV-2 |
| **Age-range** | The age-bracket (categorical) of the person, set to census data deciles from 0 to 100. Used in this simulation to capture differences in exposure risk through school closures and workforce status. |
| **Risk of death** | The overall risk of death (float) *for* each person based on their age-profile. Purely used in this simulation to remove the agents dying during the 100-day simulation time. |
| **Location** | Agents interact in over a 2-dimensional plane with their location recorded at each time-step via an x/y coordinate system. |

| | |
|---|---|
| **Span** | The distance the person moves around the environment away from their home location – longer distances result in higher likelihood of close contact with novel other people (agents) in the model. |
| **Heading / Distance** | The direction and extent of travel of the person at the current time-step. The heading and speed variables combine to create local communities and control interaction between and across communities. At higher lockdown stringency levels, agents are restricted to movement in areas closer to their home location. |
| **Contacts** | A count (integer) of contacts the person (agent) had interacted with in the past day as they moved within the model's environment. This is used in estimation of contacts with transmission potential each time-step and calculation of individual reproduction numbers at the end of infectious periods. |

Code for ABM at: https://github.com/JTHooker/COVIDModel (last accessed 23 August 2020).

ODD protocol at: https://github.com/JTHooker/COVIDModel/blob/master/ODD%20Protocol%20Aus%20NZ%20COVID19%20model.pdf (last accessed 23 August 2020).

¥ Assumed parameter based on expert opinion in conjunctions with available public data sources such as Google COVID-19 mobility reports.

¥¥ 10% of the population potentially transmit infections widely through occasional travel to random locations.

¥¥¥ The source paper reports an adjusted odds ratio of 0.15 for a systematic review of observational studies. Given possible residual confounding, and to be conservative, we used 80% rather than 85%.

*This reports all cases known to the model user on day 6 of their infection. In alternative modes, code also allows for under-reporting under extreme pressure on the track and trace system (e.g., in unmitigated scenarios).

₤ % mask wearing is fixed part of scenario, therefore no uncertainty.

#### Supplementary Table 4: Key input parameters by level of policy stringency in the ABM

| Condition | Stage 0 | Stage 1 | Stage 2 | Stage 3 | Stage 4 |
|---|---|---|---|---|---|
| % of working age adults classified as essential workers (with no restrictions on movements during work hours) *[no uncertainty, as part of scenario definition]* | 100% | 75% | 50% | 25% | 20% |
| Restrictions on non-essential workers, and essential workers when not working: | | | | | |
| % of people with restricted movement | 0% | 25% | 65% | 85% | 90% |
| % restriction in movement among the above restricted people | 0% | 25% | 65% | 85% | 90% |
| Complacency: Minimal value that restrictions above reduce to as a result of fatigue € | 0% | 15% | 52% | 68% | 81% |
| Radius of movement in spatial units for non-essential workers, and essential workers when not working † | 30 | 30 | 15 | 10 | 5 |
| Quarantine compliance ₵ – *beta distribution* | 93% | 93% | 93% | 93% | 93% |
| Super-spreader potential (generated by allowing a percentage of agents to randomly move to a new location at any time-step) | 10% | 10% | 10% | 5% | 2% |
| Limitations on gathering restrictions over time – opportunities per week to gather in locations (average of once per week) and the potential area in spatial units within which people may be drawn from. The larger the area, the greater the number of potential contacts. | 78.5 area units | 50.2 units | 28.3 units | 12.6 units | 3.1 units |
| School closures, all children < 18 years ‡ *[no uncertainty, as part of scenario definition]* | 0% | 0% | 0% | 90% | 100% |
| Mask utilisation outside of home in busy indoor environments, selected outside environments (e.g., sporting venues) and public transport where physical distancing is not possible. | 50% | 90% | 90% | 90% | 90% |
| Mask effectiveness in reducing transmission ₤ *[Beta distribution 24.3, 8.08]* | 75% | 75% | 75% | 75% | 75% |
| % of population with COVID-Safe App on their phone | 30% | 30% | 30% | 30% | 30% |
| % reduction in contact tracing time due to COVID-Safe App, when both people have the App | 50% | 50% | 50% | 50% | 50% |

† The range of movement is in a two-dimensional plane, meaning the relative difference in number of destinations is a function of the quadratic, e.g., for Stage 5 c.f. Stage 1, 25 to 4 relative difference.

‡ For this paper, all children <18 years treated the same (but can be stratified in extensions to modelling)

€ At each time-step, both the proportion of people who complied with social distancing measures and the proportion of time they complied declined by 1 unit to the baseline level set at each stage.

₤ A recent systematic review found a pooled OR for reduced transmission of 0.85 (or 85%), in mostly clinical studies and some community studies.[7] This probably overestimate effectiveness in real-life.  We therefore specified a beta distribution 24.3 and 8.08, giving mean 0.75, SD 0.075, 95% uncertainty interval 0.590 to 0.881.

**Supplementary Table 5: Input parameters to PMSLT (excluding those from ABM) and GDP costs (inputs in *italics* only used in sensitivity analyses)**

| Input | Specification | Uncertainty | Comment and source |
|---|---|---|---|
| Population counts | Estimated usually resident population 2020 | Nil | UN World Population Prospects for Jul 2020 † |
| All-cause mortality rates | Single year of age mortality rates, generated from GBD five-year age group rates using interpolation on log scale. | Log normal approximation to GBD published 2.5th and 97.5th percentiles. ‡ | IHME GHDx |
| All-cause morbidity rates | Single year of age prevalent years of life with disability (YLD) proportions | Log normal approximation to GBD published 2.5th and 97.5th percentiles. ‡ | IHME GHDx |
| *Cause-specific mortality rates (road traffic crash)* | *GBD five-year age group mortality rates.* | *(Nil – only used in sensitivity analyses as expected values)* | *IHME GHDx* |
| *Cause-specific morbidity rates (road traffic crash non-fatal injuries, depression, anxiety)* | *Single year of age prevalent years of life with disability (YLD) proportions for these for conditions.* | *(Nil – only used in sensitivity analyses as expected values)* | *IHME GHDx* |
| Forecast annual percentage change (APC) in all-cause mortality rates | APC by sex by five-years age-groups for GBD mortality rates 1980-2017, used to forecast mortality rates to 2035 – then no change. | Nil | IHME GHDx |
| Total health system expenditure per person by sex and age | Average per-capita health expenditure by sex and age group. | 5% SD log-normal distribution. ‡ | Australia: AIHW [8] |
| *Policy strategy (i.e., intervention) inputs* | | | |
| SARS-CoV-2 mortality | Varies with SARS-CoV-2 infection rates and specified in detail in Appendix 3. | See Appendix 3. ‡ | See Appendix 3. |
| SARS-CoV-2 morbidity | Symptomatic, not admitted = 0.084 | 95% UI 0.059 – 0.110 | See Appendix 3. |
| | Admitted to hospital, no ICU = 0.096 | 95% UI 0.064 – 0.128 | |
| | Admitted to ICU = 0.283 | 95% UI 0.208 – 0.359 | |
| SARS-CoV-2 health expenditure | Symptomatic, not admitted = US$57.68 | +/- 20% SD log normal distribution | See Appendix 3. |
| | Admitted to hospital, no ICU = US$14,324 | | |
| | Admitted to ICU =US$44,641 | | |
| GDP impacts | Varies by stage:<br>- US$0.4 billion/ week Stage 1 | +/- 20% SD log normal distribution. Correlated 1.0 across timesteps. | See Appendix 1. |

| Input | Specification | Uncertainty | Comment and source |
|---|---|---|---|
| | - US$0.6 billion/ week Stage 1b<br>- US$0.725 billion/ week Stage 2<br>- US$1.275 billion/ week Stage 3<br>- US$2.61 billion/ week Stage 4 | | |
| *Road traffic crash (RTC) rates* | *The correspondence to changes in Apple mobility data to changes in Victoria RTC rates was used to generate the following estimates: -23.6%, -21.9%, -8.74%, -1.53%, -1.45% reductions for stages 4, 3, 2, 1b and 1a, respectively.* | *(Nil – only used in sensitivity analyses as expected values)* | *See Appendix 4 for details.* |
| *Depression and anxiety* | *We assumed the following percentage increases in both depression and anxiety: 10%, 8%, 6%, 4% and 2% for stages 4, 3, 2, 1b and 1a, respectively.* | *(Nil – only used in sensitivity analyses as expected values)* | *See main paper for details.* |

Footnotes:

Values for Australia were applied to Victoria (except population being scaled).

Abbreviations: AIHW = Australian Institute of Health and Welfare; GHDx = Global Health Data Exchange; IHME = Institute of Health Metrics and Evaluation; (http://ghdx.healthdata.org/);

† https://population.un.org/wpp/Download/Standard/Population/

‡ Each set of variables, for instance, variations in infection fatality rates by age and sex, have a 0.5 correlation within each iteration. All variables assumed to be correlated perfectly (1.0) across time-steps within each iteration.

**Supplementary Table 6: Sensitivity analyses incremental health loss (HALYs) compared to BAU (i.e., no SARS-CoV-2 pandemic) and additional health expenditure and GDP loss (3% discount rate; in US$ millions), using the median infection rate across 100 simulations (%in parentheses are change relative to baseline)**

| Sensitivity analysis | Measure (lifetime) | a) Aggressive elimination | b) Moderate elimination | c) Tight Suppression | d) Loose suppression |
|---|---|---|---|---|---|
| Baseline † | HALY loss | 289 | 308 | 2,080 | 12,700 |
| | Net health Costs | -3.13 | -3.31 | -21.8 | -131 |
| | Net health + GDP costs | 46,381 | 41,658 | 50,808 | 50,235 |
| | *Optimal from health perspective* | *> $10,000* | | | *<$10,000* |
| | *Optimal from partial societal perspective* | | *$0 to $500,000* | | |
| *Discount rate* | | | | | |
| 0% | HALY loss | 367 (27%) | 391 (27%) | 2,670 (28%) | 16,500 (30%) |
| | Net health Costs | -4.94 (58%) | -5.24 (58%) | -35.2 (61%) | -215 (64%) |
| | Net health + GDP costs | 46,378 (0%) | 41,655 (0%) | 50,792 (0%) | 50,171 (0%) |
| | *Optimal from health perspective* | *> $15,000* | | | *<$15,000* |
| | *Optimal from partial societal perspective* | | *$0 to $500,000* | | |
| 6% | HALY loss | 239 (-17%) | 253 (-18%) | 1,690 (-19%) | 10,300 (-19%) |
| | Net health Costs | -1.97 (-37%) | -2.07 (-37%) | --13.3 (-39%) | -78.3 (-40%) |
| | Net health + GDP costs | 46,383 (0%) | 41,660 (0%) | 50,817 (0%) | 50,275 (0%) |
| | *Optimal from health perspective* | *> $10,000* | | | *<$10,000* |
| | *Optimal from partial societal perspective* | | *$0 to $500,000* | | |
| *Varying timeline to vaccine (from 12 months in base model)* | | | | | |
| 6 months | HALY loss | 283 (-2%) | 300 (-3%) | 1,050 (-50%) | 4,340 (-67%) |
| | Net health Costs | -3.07 (-2%) | -3.25 (-2%) | -11.2 (-49%) | -45.8 (-65%) |
| | Net health + GDP costs | 30,419 (-34%) | 28,604 (-31%) | 26,978 (-47%) | 25,033 (-50%) |
| | *Optimal from health perspective* | *> $10,000* | | | *<$10,000* |
| | *Optimal from partial societal perspective* | | | | *$0 to $500,000* |
| 18 months | HALY loss | 291 (1%) | 312 (1%) | 3,000 (44%) | 15,000 (18%) |
| | Net health Costs | -3.15 (1%) | -3.36 (2%) | -30.9 (42%) | -154 (18%) |
| | Net health + GDP costs | 61,233 (32%) | 56,723 (36%) | 71,328 (40%) | 67,716 (35%) |
| | *Optimal from health perspective* | *> $10,000* | | | *<$10,000* |
| | *Optimal from partial societal perspective* | | *$0 to $500,000* | | |

| **Sensitivity analysis** | **Measure (lifetime)** | **a) Aggressive elimination** | **b) Moderate elimination** | **c) Tight Suppression** | **d) Loose suppression** |
|---|---|---|---|---|---|
| *Including additional diseases* | | | | | |
| Road traffic crash (RTC) | HALY loss | -661 (-329%) | -684 (-322%) | 914 (-56%) | 11,900 (-6%) |
| | Net health Costs | 4.99 (-259%) | 5.18 (-256%) | -11.8 (-46%) | -123 (-6%) |
| | Net health + GDP costs | 46,391 (0%) | 41,668 (0%) | 50,820 (0%) | 50,243 (0%) |
| | Optimal from health perspective | | > $10,000 | | <$10,000 |
| | Optimal from partial societal perspective | | $0 to $500,000 | | |
| Depression and anxiety | HALY loss | 738 (155%) | 762 (147%) | 2,620 (26%) | 13,200 (4%) |
| | Net health Costs | -3.13 (0%) | -3.31 (0%) | -21.8 (0%) | -131 (0%) |
| | Net health + GDP costs | 46,381 (0%) | 41,658 (0%) | 50,808 (0%) | 50,235 (0%) |
| | Optimal from health perspective | > $10,000 | | | <$10,000 |
| | Optimal from partial societal perspective | | $0 to $500,000 | | |
| RTC, depression & anxiety | HALY loss | -212 (-173%) | -229 (-174%) | 1,460 (-30%) | 12,300 (-3%) |
| | Net health Costs | 4.99 (-259%) | 5.18 (-256%) | -11.8 (-46%) | -123 (-6%) |
| | Net health + GDP costs | 46,391 (0%) | 41,668 (0%) | 50,820 (0%) | 50,243 (0%) |
| | Optimal from health perspective | | > $10,000 | | <$10,000 |
| | Optimal from partial societal perspective | | $0 to $500,000 | | |
| *Contact tracing (effectiveness improves inversely with the log of daily cases)* | | | | | |
| | HALY loss | 317 (10%) | 332 (8%) | 3,470 (67%) | 14,300 (13%) |
| | Net health Costs | -3.43 (10%) | -3.59 (8%) | -35.8 (64%) | -148 (13%) |
| | Net health + GDP costs | 47,766 (3%) | 44,356 (6%) | 53,845 (6%) | 54,548 (9%) |
| | Optimal from health perspective | > $10,000 | | | <$10,000 |
| | Optimal from partial societal perspective | | $0 to $500,000 | | |

**Supplementary Figure 1: Stages and triggers for tight suppression**

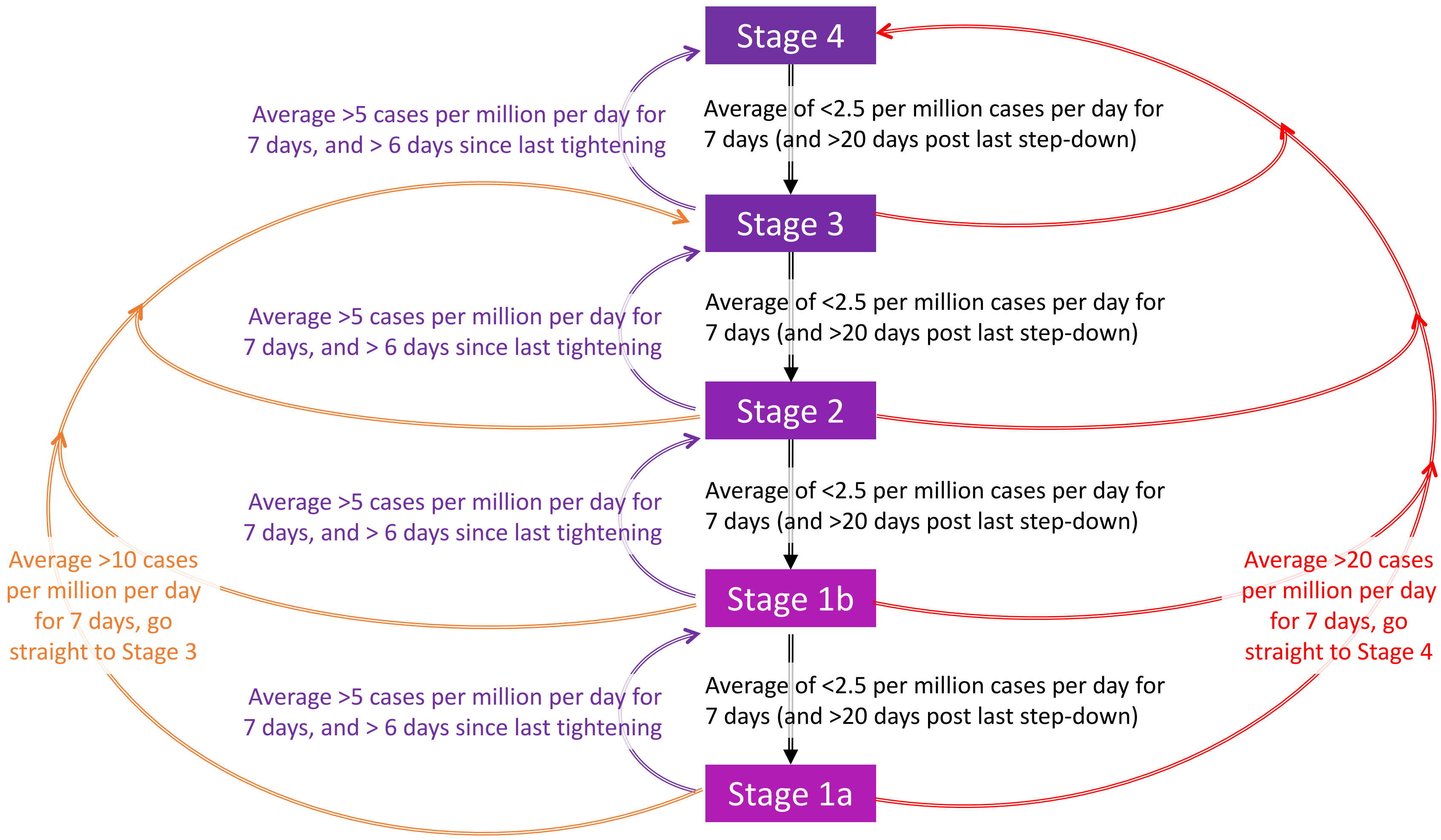

**Supplementary Figure 2: Net monetary benefit ($US) at per capita GDP ($US 55,000) using median expected values:**

#### a) Health system perspective

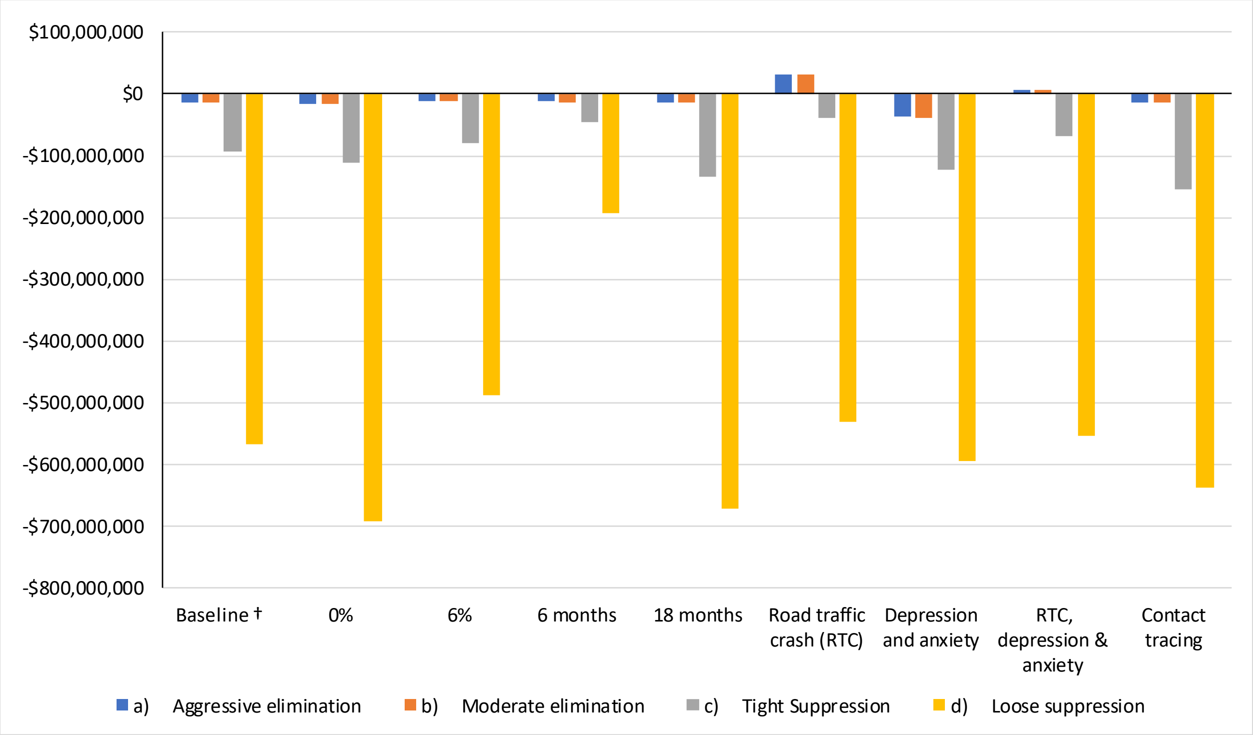

#### a) Partial societal perspective

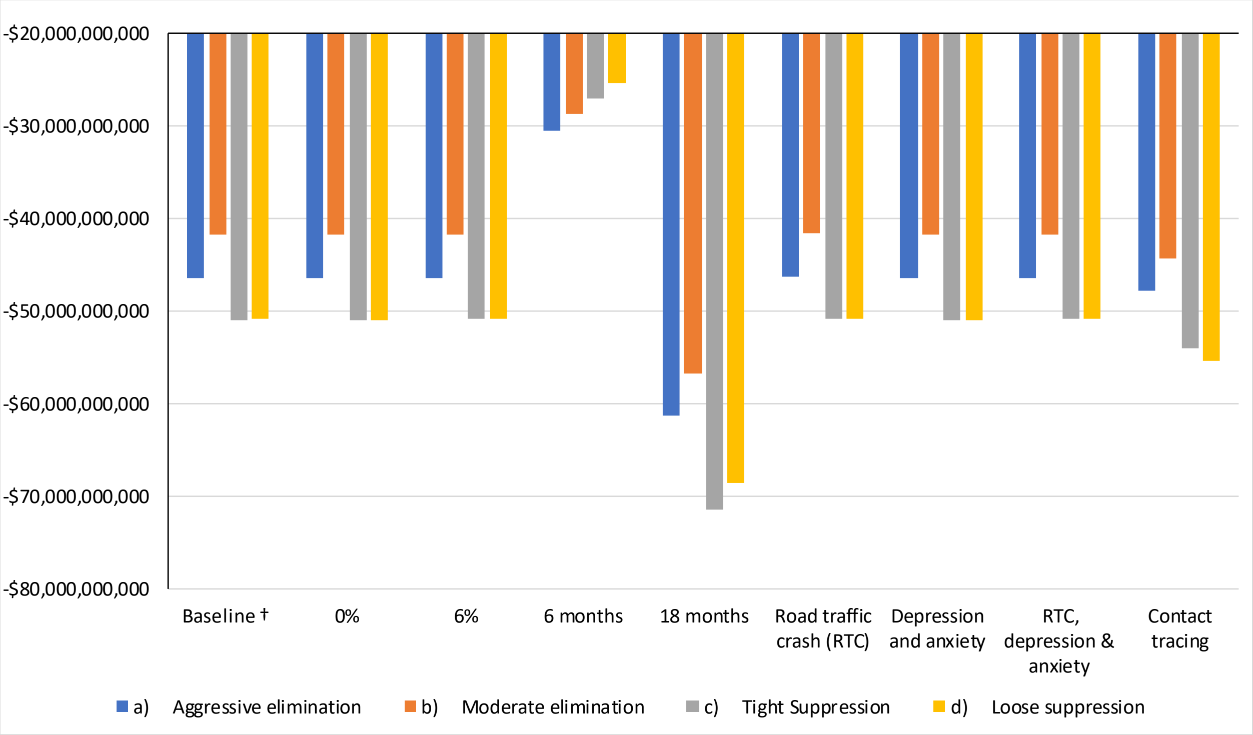

#### Appendix 1 Estimates of GDP loss by stage, from Australian and Victorian Treasuries

State and Commonwealth treasuries in Australia provided estimates of the impact of COVID-19. We prioritised the use of such government estimates since these are usually bolstered by a wide array of near real-time indicators, for instance, income and sales tax collection data.

The Victorian Treasury in its July 2020 update, before the announcement of Stage 4 restrictions, estimated that a combination of approximately six weeks of Stage 3 and six weeks of various Stage 2 lockdowns in the April to June and July to September quarters would reduce GDP by 11 per cent (or roughly 1 billion dollars a week compared to expected GDP had there been no COVID-19).[9]

For each stage, the broader Australian economy had a smaller impact from equivalent restrictions, in part due to a smaller reliance outside of the State of Victoria on hospitality and the resilience of iron ore prices. The Australian Department of Finance in its Mid-Year Economic and Financial Update estimated that Stage 3 restrictions between Mar 30 and mid-May had an estimated Australia-wide cost of AUD 4 billion per week, around, roughly 11 per cent of the weekly Australian GDP of AUD 36.3 billion dollars (~AUD1.89 trillion/52). It estimated the increment between Stage 3 and the 'unlocked' economy to be around AUD 2bn per week or around 5.5% of GDP. Finally, the prime minister of Australia announced on 6 Aug 2020 [10], that the cost of six weeks of Victorian Stage 3 restrictions in the Jul-Sep quarter would be around AUD 3.3 billion, and the cost of six weeks of Stage 4 restrictions, incremental to Stage 3, was AUD 7 to 9 billion. Approximately 80 per cent, or $6 billion to $7 billion, was expected to arise from businesses and activity in Victoria, while the remainder cost was borne by the rest of Australia due to spill-over effects.

Based on these announcements and using a scope of including GDP losses caused by Victoria even if borne beyond Victoria, we estimate the impact of the COVID-19 control strategies to be approximately as shown below in Supplementary Table 7.

**Supplementary Table 7: Estimates of per week GDP loss by stage, relative to no restrictions. Scope included GDP losses caused by Victorian restrictions, but borne beyond Victoria**

| *Stage* | Total GDP costs for each stage per week, relative to no restrictions (i.e. pre-COVID; AUD b,) | Incremental GDP costs for each higher stage, per week (AUD bn) | Rationale |
|---|---|---|---|
| *Stage 1* | $0.5 | -- | 2 bn/week cost of most relaxed Stage estimated for all of Australia [11]; Victoria ~ 0.25% of Australian economy. |
| *Stage 1b* | $0.535 | $0.035 | Includes incremental 150 million monthly turnover impact based on 70% turnover growth observed in NSW between May and August. [12]<br>(Minimal impact on take-away services. Minimal incremental impact on real estate auctions.) |
| *Stage 2* | $0.725 | $0.19 | Backed out from incremental cost of Stage 3 over Stage 2 by Australian Treasury; VIC treasury forecasts of combined Stage 2-Stage 3 impacts.[9]<br>0.5x+0.5(x+0.55) =1<br>→ x + 0.275 = 1 |

| *Stage* | **Total GDP costs for each stage per week, relative to no restrictions (i.e. pre-COVID; AUD b,)** | **Incremental GDP costs for each higher stage, per week (AUD bn)** | **Rationale** |
|---|---|---|---|
| | | | → x = 0.725 |
| *Stage 3* | $1.275 | $0.55 | Prime Minister announcement/Australian Treasury's calculations of an incremental (compared to Stage 2) 0.55 billion costs per week for around 6 weeks of Stage 3 restrictions in VIC. [10], |
| *Stage 4* | 2.61 (2.45-2.78) | 1.33 (1.16-1.5) | Incremental cost of Stage 4 compared to Stage 3. Incremental costs of 7-9 billion over six weeks. [10], |

Note: A 20% SD was assumed for all these inputs, to draw values from in Monte Carlo estimates to simulate uncertainty around outputs from modelling.

#### Appendix 2 Average citizen annual health expenditure

Consistent with recommended practice in cost effectiveness analyses [13,14], in the USA [15] and the Netherlands[16], we included 'unrelated disease costs' in the economic evaluation. This means that in addition to including the costs of SARS-CoV-2 cases per se (Appendix 3), knock-on changes in health system expenditure are also included. For SARS-CoV-2, this means that if someone dies due to SARS-CoV-2 infections, their reduced health expenditure in the future is included (leading to a potentially net negative expenditure depending on the balance of costs, age and discount rate). In a simulation model, this is easy to incorporate, by including an expenditure reward per cycle in the model for diseases not explicitly modelled elsewhere – which in the case of SARS-CoV-2 modelling, is simply the expected annual (or monthly) average health system expenditure.

Data were extracted from the Australian Institute of Health and Welfare (AIHW) report 'Disease expenditure in Australia, which separates the total expenditure by sex and age.[8]

The data are from the 2015-16 financial year, where the total health expenditure totalled $170.4 billion $AU (2016). The AIHW attributed $106.857 billion of this spending to age and sex related health spending (62.7% of total health expenditure), with data provided as total expenditure by age and sex subgroup. [17]

We extracted population demographics from the Australian Bureau of Statistics (ABS) 2016 population report,[17] and the total health expenditure for each subgroup was then divided by the corresponding population numbers for these subgroups. Thus the 2016 health expenditure is expressed as per capita expenditure, by age and sex.[18]

The AIHW estimates variable health expenditure at 94% of total health expenditure,[8] whilst New Zealand variable expenditure is estimated at 91% total expenditure.[19] We elected to assume variable expenditure was 90%, allowing for fixed costs in running services Noting the above 62.7% of total health expenditure captured by AIHW estimates, we therefore multiplied all age by sex empirical estimates by a factor of 90/62.7 to generate the estimated predicted Australian variable health expenditure per capita, by age and sex.

Next, we inflation adjusted these expenditures from 2016 $AU to 2019 $AU using Australian CPI adjustment factors (OECD rates; [18] https://data.oecd.org/price/inflation-cpi.htm). Finally, we converted to 2019 USD using the AUD-USD 2019 purchasing power from the OECD.

**Supplementary Table 8: Per-capita annual health expenditure in Australia, by sex by age (2019 US$)**

| Age | Male | Female |
|---|---|---|
| **<1** | 9537.35 | 8225.14 |
| **1-4 years** | 1946.15 | 1540.07 |
| **5-9 years** | 1407.47 | 1158.21 |
| **10-14 years** | 1357.51 | 1278.25 |
| **15-19 years** | 1677.19 | 2248.60 |
| **20-24 years** | 1710.49 | 2897.45 |
| **25-29 years** | 1823.23 | 3808.70 |
| **30-34 years** | 2110.59 | 4848.75 |
| **35-39 years** | 2521.40 | 4582.59 |
| **40-44 years** | 3040.05 | 4113.00 |
| **45-49 years** | 3465.57 | 4041.90 |
| **50-54 years** | 4489.94 | 4757.85 |

| Age | Male | Female |
|---|---|---|
| **55-59 years** | 5612.20 | 5344.07 |
| **60-64 years** | 7175.25 | 6410.48 |
| **65-69 years** | 9246.18 | 8091.24 |
| **70-74 years** | 11366.34 | 9822.93 |
| **75-79 years** | 16516.38 | 12533.93 |
| **80-84 years** | 17037.19 | 14161.53 |
| **85+ years** | 19049.62 | 15568.81 |

#### Appendix 3 SARS-CoV-2 parameters

For each monthly cycle, the number of SARS-CoV-2 infections were split into the following categories for all modelled cases, equivalent to all notified and confirmed cases[1]:

A. Asymptomatic
B. Symptomatic, not admitted to hospital
C. Symptomatic, admitted to hospital
D. Symptomatic, admitted to hospital and ICU
E. Die (may come from anyone of B, C and D)

This is slightly different from our previous model [20] as there is now sufficient within-Australia data (i.e. for Victoria, from the Victorian Department of Health and Human Services (Vic DHHS)) to estimate probabilities of hospitalisation, ICU admission and death directly. A large fraction of people dying did not get admitted to ICU, dying on a general ward or in community care – especially elderly people with a do not resuscitate order. We therefore estimated proportions of cases into four mutually exclusive categories (A, B, C and D) for the quantification of morbidity and health expenditure, and one additional category for the quantification of HALYs lost due to death (E).

In this Appendix we describe in order:

- The epidemiological parameters to split each month's SARS-CoV-2 infections into the five above categories.
- The excess health expenditure assigned to each of the three symptomatic SARS-CoV-2 categories (B, C and D).
- The morbidity-loss assigned to each of the three symptomatic SARS-CoV-2 categories (B, C and D).

#### Epidemiological parametrisation

Supplementary Table 9 (below) shows the number of cases, hospitalisations, ICU admissions and deaths in Victoria. The dates are deliberately different, so that average time lags are allowed for: up to 14 days from notification to death; subsuming 10 days from notification to ICU admission; subsuming 7 days from notification to hospitalisation.

[1] Undetected cases are not modelled explicitly. They exist (to an unknown degree) but assumed to not incur (much) morbidity or cost.

##### Supplementary Table 9: Numbers of confirmed SARS-CoV-2 cases, hospitalisations, ICU admission and deaths in Victoria, Australia

| Sex* | Age | Confirmed cases in VIC: 1 Jan to 14 Aug | Hospitalisations (for cases diagnosed between 1 Jan to 20 Aug) | ICU admissions (for cases diagnosed between 1 Jan and 23 Aug) | Deaths (for cases diagnosed between 1 Jan and 14 Aug) |
|---|---|---|---|---|---|
| Female | 0-9 | 434 | 14 | 0 | |
| | 10-19 | 774 | 9 | 1 | |
| | 20-29 | 1,987 | 74 | 8 | 0 |
| | 30-39 | 1,445 | 83 | 4 | 0 |
| | 40-49 | 1,082 | 81 | 13 | 0 |
| | 50-59 | 975 | 110 | 21 | 3 |
| | 60-69 | 542 | 122 | 20 | 5 |
| | 70-79 | 337 | 162 | 23 | 26 |
| | 80-89 | 534 | 309 | 5 | 112 |
| | 90+ | 379 | 188 | 1 | 86 |
| Male | 0-9 | 479 | 13 | 2 | |
| | 10-19 | 879 | 7 | 1 | |
| | 20-29 | 1,904 | 42 | 3 | 1 |
| | 30-39 | 1,458 | 54 | 7 | 2 |
| | 40-49 | 1,015 | 99 | 15 | 1 |
| | 50-59 | 847 | 141 | 36 | 9 |
| | 60-69 | 518 | 159 | 32 | 13 |
| | 70-79 | 375 | 179 | 21 | 53 |
| | 80-89 | 306 | 198 | 8 | 97 |
| | 90+ | 154 | 102 | 0 | 67 |
| Total | | 16424 | 2146 | 221 | 475 |

Supplementary Figure 3 shows the ln odds of hospitalisation, ICU admission and deaths for observed data when the number of events is 5 or more, and from simple predictive logistic regression models on the same data. For the latter regressions, main effects were included for sex and age as a continuous variable, and additional age-dummies due to non-linearity on the ln odds scale for:

- Hospitalisation: 0-9, 10-19, 80-89 and 90+ year olds
- ICU admission: 80-89 and 90+ year olds
- Deaths: 90+ year olds.

**Supplementary Figure 3: Ln odds of hospitalisation, ICU admission and death for confirmed cases for observed data when number of events > 5 and from a logistic regression prediction otherwise**

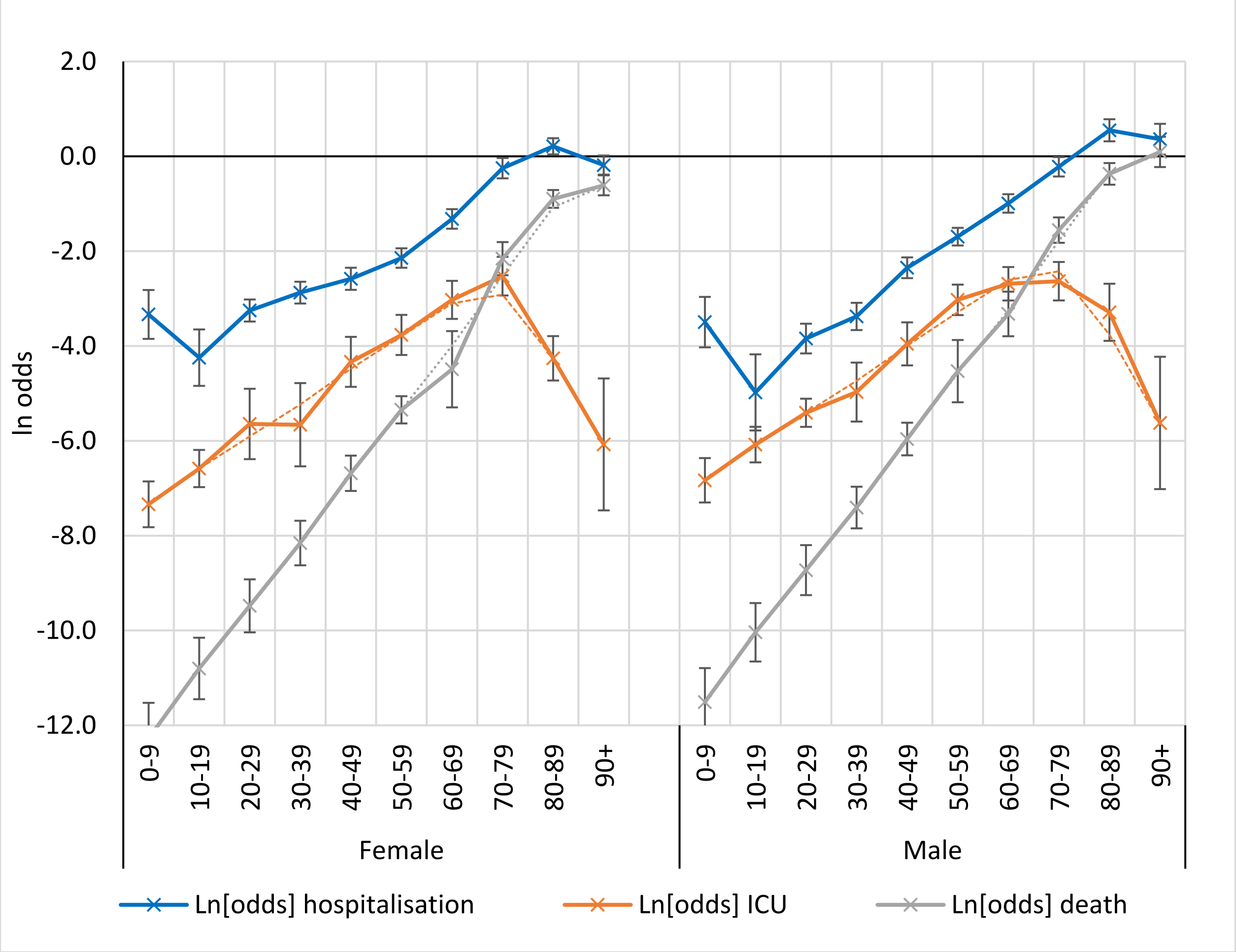

Error bars are 95% confidence intervals

We elected to use the observed ln odds when the number of events was five or more, otherwise use the logistic regression predicted ln odds. These estimates, and their standard errors, are shown in Supplementary Table 10.

Sequentially, the process to estimate the actual disaggregation of SARS-CoV-2 cases (outputted by the ABM) by category of morbidity and mortality was:

- Estimate the monthly number of deaths (E) by sex and age; propagate through the PMSLT increasing mortality rates (no change in morbidity)
- Estimate the number of ICU admissions (D) by sex and age; propagate through the PMSLT increasing morbidity and health expenditure (no change in mortality)
- Estimate the number of hospitalisation admissions (B) by sex and age, subtracting off ICU admissions; propagate through the PMSLT increase morbidity and health expenditure (no change in mortality)
- Estimate number of asymptomatic cases (A).

- And finally, estimate the number of symptomatic cases, as the total number of cases, minus (A + C + D + E). And link them to morbidity and health expenditure.

(The estimates of morbidity and excess health expenditure are described in subsequent sections of this Appendix.)

**Supplementary Table 10: ln odds (standard errors) used in simulation modelling**

| | | Hospitalisation | | ICU | | Death | |
|---|---|---|---|---|---|---|---|
| **Sex** | **Age** | **ln(odds)** | **s.e. ln(odds)** | **ln(odds)** | **s.e. ln(odds)** | **ln(odds)** | **s.e. ln(odds)** |
| Female | 0-9 | -3.335 | 0.263 | -7.339 | 0.246 | -12.276 | 0.383 |
| | 10-19 | -4.246 | 0.304 | -6.584 | 0.200 | -10.800 | 0.331 |
| | 20-29 | -3.252 | 0.118 | -5.644 | 0.379 | -9.481 | 0.285 |
| | 30-39 | -2.874 | 0.117 | -5.660 | 0.448 | -8.156 | 0.240 |
| | 40-49 | -2.583 | 0.119 | -4.334 | 0.269 | -6.685 | 0.190 |
| | 50-59 | -2.145 | 0.105 | -3.766 | 0.216 | -5.346 | 0.147 |
| | 60-69 | -1.321 | 0.105 | -3.027 | 0.205 | -4.491 | 0.411 |
| | 70-79 | -0.250 | 0.110 | -2.527 | 0.208 | -2.158 | 0.179 |
| | 80-89 | 0.211 | 0.087 | -4.260 | 0.239 | -0.898 | 0.096 |
| | 90+ | -0.184 | 0.103 | -6.076 | 0.710 | -0.612 | 0.107 |
| Male | 0-9 | -3.497 | 0.271 | -6.832 | 0.239 | -11.510 | 0.366 |
| | 10-19 | -4.979 | 0.410 | -6.079 | 0.191 | -10.038 | 0.315 |
| | 20-29 | -3.843 | 0.160 | -5.408 | 0.151 | -8.727 | 0.269 |
| | 30-39 | -3.377 | 0.147 | -4.972 | 0.317 | -7.407 | 0.224 |
| | 40-49 | -2.350 | 0.112 | -3.955 | 0.232 | -5.962 | 0.176 |
| | 50-59 | -1.694 | 0.095 | -3.027 | 0.164 | -4.530 | 0.335 |
| | 60-69 | -0.993 | 0.099 | -2.688 | 0.180 | -3.324 | 0.240 |
| | 70-79 | -0.221 | 0.104 | -2.633 | 0.207 | -1.556 | 0.136 |
| | 80-89 | 0.549 | 0.119 | -3.289 | 0.307 | -0.370 | 0.116 |
| | 90+ | 0.362 | 0.165 | -5.623 | 0.712 | 0.093 | 0.163 |

**Supplementary Table 11: Parametrisation of SARS-CoV-2 asymptomatic cases**

| Age Group | Percentage symptomatic, with 20% of all infections across age asymptomatic | | |
|---|---|---|---|
| | Percentage symptomatic | Mean (ln odds scale) | SD |
| 0 to 9 | 35% | -0.639 | 0.302 |
| 10 to 19 | 56% | 0.235 | 0.302 |
| 20 to 29 | 75% | 1.109 | 0.302 |
| 30 to 39 | 88% | 1.983 | 0.302 |
| 40 to 49 | 95% | 2.857 | 0.302 |
| 50 to 59 | 98% | 3.731 | 0.302 |
| 60 to 69 | 99% | 4.605 | 0.302 |
| 70 to 79 | 100% | 5.479 | 0.302 |
| 80+ | 100% | 6.353 | 0.302 |

#### Morbidity loss by SARS-CoV-2 category

An 'average' incremental morbidity impact due to SARS-CoV-2 for each month was estimated as the weighted sum of the morbidity impact for each of the five symptomatic SARS-CoV-2 categories (weighted by proportionate distribution of SARS-CoV-2 infections by category – which varied iteration to iteration given the uncertainty described above). We measured morbidity impacts using disability rates (DR),[21] according to severity of acute infection (mild, moderate and severe). For ICU admissions, DR were based on severe chronic obstructive pulmonary disease (COPD) to reflect Acute Respiratory Distress Syndrome (ARDS). We assume all survivors return to their baseline health status (pre-SARS-CoV-2) (DR: 0) following a specified recovery period as described below.

Morbidity loss for the four categories of symptomatic SARS-CoV-2 infection include:

- Morbidity for people admitted to ICU, but surviving, assuming a mean duration from symptom onset to recovery of 6 weeks was based on the higher range of the median time from onset to clinical recovery for patients with severe or critical disease.[22] We applied a DR for moderate acute infectious episode for 1 week of 0·051 (0·032–0·074), plus DR for severe acute infection for 2 weeks of 0·133 (0·088–0·190), and ICU admission for 1 week of 0.408 (0.273-0.556), plus a return to baseline health (DR: 0) over 2 weeks (equivalent to 50% probability of ARDS for 2 weeks).
- Morbidity for people admitted to hospital but not requiring ICU, assuming a mean duration from symptom onset to recovery of 4 weeks, based on the lower range of the median time from onset to clinical recovery for patients with severe or critical disease. We applied a DR for moderate acute infectious episode for 1 week of 0·051 (0·032–0·074), plus severe infectious episode for 2 weeks of 0·133 (0·088–0·190), plus return to baseline health (DR: 0) over 1 week (equivalent to 50% probability of severe infectious episode for one week).
- Morbidity for people diagnosed with symptomatic disease but not admitted to hospital, assuming a mean duration from symptom onset to recovery of 2.5 weeks, based on data from the WHO-China Joint Mission report on median time from onset to clinical recovery for mild cases of approximately 2 weeks.[23] We assume half of people with symptomatic disease who are not admitted to hospital have mild symptoms, and the other half have moderate symptoms.[22] Therefore we applied a DR of 0·051 (0·032–0·074) for moderate acute infection for 50% of this group for 2.5 weeks and a DR of 0·006 (0·002–0·012) for mild acute infection for 2.5 weeks for the remaining 50% of this category.

The overall morbidity for each SARS-CoV-2 category was calculated as the sum of the symptom severity specific DR multiplied by symptom duration.

**Supplementary Table 12: Disability rate distribution and duration by SARS-CoV-2 category**

| Category | Symptom severity (health state) | Duration | Disability rate, Mean (95% CI) |
|---|---|---|---|
| Admitted to ICU, | Moderate acute infection | 1 week | 0.051 (0.032-0.074) |
| | Severe acute infection | 2 weeks | 0·133 (0·088–0·190) |
| | In-ICU (COPD for ARDS) | 1 weeks | 0.408 (0.273-0.556) |
| | Return to baseline health | 2 weeks | Linearly to 0 from ARDS DR |
| Admitted to hospital, no ICU | Moderate acute infection | 1 week | 0.051 (0.032-0.074) |
| | Severe acute infection | 2 weeks | 0.133 (0·088–0·190) |
| | Return to baseline health | 1 week | Linearly to 0 from severe acute infection DR |
| Symptomatic, not admitted | 50% Moderate acute infection | 2.5 weeks | 0.051 (0.032-0.074) |
| | 50% Mild acute infection | 2.5 weeks | 0·006 (0·002–0·012) |

We assumed those who died from SARS-CoV-2 in the community have the same morbidity as ICU deaths.

#### Excess health expenditure by SARS-CoV-2 category

We estimated the excess health expenditure by SARS-CoV-2 category using an ingredients approach. For each patient category, we modelled the expected patient pathway through the health system, based on available SARS-CoV-2 data from China, Australia, and the UK (Supplementary Tables below). For each patient subgroup category described in the paper, we calculated total health expenditure by first estimating resource use required for a typical patient (e.g., hospital or outpatient visits, or drugs), and multiplying by unit costs for each of the specified resources.

Supplementary Table 13 shows resource use assumptions based on clinical costing data. All patient categories are assumed to have similar patient pathways. Patients who die can come from several of the patient categories below. Current Victorian data has shown that many elderly patients who died as a result of SARS-CoV-2 infection did not receive ICU care. Unfortunately, the data did not allow us to determine exactly what proportion of patients who died accessed ICU care. From the available data, which captures all Victorian SARS-CoV-2 patients up to August 28, we determined that if all possible deaths came from ICU wards, 18% of all deaths (94/513) would have come from an ICU ward. We know that all ICU-admissions will not result in death. We have therefore conservatively estimated that 10% of all patients who die from SARS-CoV-2, have incurred an ICU visit. While it is understood that most COVID-19 deaths in Victoria have occurred in hospital, some deaths have occurred in aged care facilities without being transferred to hospital.[24] As it currently unknown how many patients have died without receiving hospital treatment, we assume that end-of-life care within these facilities will require similar levels of health care as hospitalisation.

The SARS-CoV-2 pandemic is expected to add additional costs to hospital operations, adjusting for complexity of patients and added infection control required including the need for isolation of patients, staff time for proper fitting of personal protective equipment, and enhanced cleaning regiments. As a result, inpatient and ICU hospital costs have been scaled up by 20% to account for these extra costs. This 20% estimate is based on the Coronavirus Aid, Relief and Economic Security (CARES) Act in the United

States that provides a 20% add-on payment for COVID-19 patients.[25] We expect that this loading will be a moderate estimate, and likely underestimate the true hospital costs during a pandemic outbreak. [25] Total costs for each patient category described above is found in Supplementary Table 13.

##### Supplementary Table 13*:* Resource use assumptions by SARS-CoV-2 category

| Patient Subgroup | Treatment items | Source |
|---|---|---|
| **Treated in ICU and survived** | GP Visit<br><br>ER visit<br><br>ICU<br><br>Inpatient (non-ICU)<br><br>Pandemic loading | Assumption that half these patients will have contact with a GP prior to hospitalisation<br>Base fee charged for presenting to an ER<br><br>Cost of ICU day X LOS<br><br>Cost of inpatient day X LOS<br>All hospital costs have 20% loading for pandemic† |
| **Non-ICU hospitalisation and survived** | GP Visit<br><br>ER visit<br><br>Inpatient<br><br>Pandemic loading | Assume all these patients will have contact with GP prior to hospitalisation<br>Base fee charged for presenting to an ER<br><br>Cost of inpatient day X LOS<br>All hospital costs have 20% loading for pandemic |
| **Symptomatic case, no hospitalisation and survived** | GP Visit<br><br>Paracetamol | Assume patients have two GP appointments on average (initial visit and follow up appointment)<br>Assume on average all patients will purchase 1 round of paracetamol for symptom reprieve |
| **Infected case with no symptoms** | No resources | |
| **Died** | GP Visit<br><br>ER visit<br>ICU<br><br><br>Inpatient (non-ICU)<br>Pandemic Loading | Assumption that these patients will have 2 contacts with a GP prior to hospitalisation<br><br>Base fee charged for presenting to an ER<br>10% of patients in this category: Cost of ICU day X length of stay (LOS) +Cost of inpatient day X inpatient LOS<br>90% of patients in this category: Cost of inpatient day X LOS<br>All hospital costs have 20% loading for pandemic |

† https://revcycleintelligence.com/news/how-much-will-the-covid-19-pandemic-cost-hospitals; accessed 5 May 2020
ER= Emergency Room visit, ICU= Intensive care unit, LOS= Length of stay, GP= General Practitioner

Supplementary Table 14 below shows the input quantities for hospital length of stay across the different patient group categories.

**Supplementary Table 14: Quantity inputs for Hospital (inpatient and ICU) length of stay**

| Patient Subgroup | Length of Stay | Detail |
|---|---|---|
| **Died:** | | Can come from ICU, Hospital, or other. |
| **Inpatient 11 days** (90%) | 11 days (total)[26] N=8 | 90% of patients who die incur no ICU stay: FluCAN estimates people who died from COVID had a median inpatient LOS of 11 days.[26] |
| **ICU 7 days + 4 days inpatient days** (10%) | 8 days.[27]N=54, IQR (4-12)<br>6 days. [28]N=698, IQR (3-9) | Patients who died that entered ICU conservatively estimated at 10%: Median length of ICU days for patients who did not survive. Taken as an estimate between the two data points. Estimate fits within both IQRs. Assume 4 inpatient days on top of ICU LOS (from 11-day FLUCAN estimate). |
| **ICU Survived:** | | |
| **ICU 5.5 days** | 7 days. [27]N=137, IQR (2-9)<br>4 days).[28]N=355, IQR (2-8) | Median length of ICU days for patients who survived. Taken as an estimate between the two data points. Estimate fits within both IQRs. |
| **Inpatient 12 days** | 12 days.[28]N= 137, IQR (9-15) | Median length inpatient stays for patients who attended ICU and survived. Single point estimate. |
| **Non-ICU hospitalisation and survived:** | | |
| **Inpatient 11 days** | 11 days. [29]N= 926, IQR (10-13) | Median length inpatient stays for non-severe patients, not needing ICU. Single point estimate. |

* Representative of patients up until 5th July 2019. Data was not been updated on LOS for patients who have died, thus may not adequately reflect aged care outbreaks.

Unit costs for each type of health system utilisation (GP visit, emergency room (ER) visit, inpatient bed day, ICU bed day) were estimated from sources in Supplementary Table 15 (below).

##### Supplementary Table 15: Unit costs for healthcare resource use (2019 AUD)

| Resource | Source | Detail |
|---|---|---|
| Paracetamol<br><br>$6.60 | PBS item 10582Y (100 units, 500mg) | Mild cases have symptoms for 2 weeks.[30] Paracetamol recommendation for mild cases. [31] |
| GP<br>$38.20 | MBS item 23. Level B General consultation. Range ($17.50 - $108.85) | MBS telehealth items price match (3, 23, 36, 44 and 91790, 91800, 91802). Range estimates adapted to 2019 values. [32,33] |
| ER visit<br>$100.38 | Data from Manual of Resource Items and Their Associated Costs (Department of Health) [33] | 2005 price adjusted to 2019 price. Cost applied for all patients requiring hospitalisation. |
| Inpatient day $1551 | Data from H1N1 outbreak using AR-DRG code. [34] | Total cost of inpatient day post ICU. Also assumed for all non-ICU patients. Adjusted to 2019 value. |
| ICU day<br>$6331 | Micro-costed from H1N1 outbreak. [34] | Total cost per ICU bed-day. Allied health and overheads included. Adjusted to 2019 value |

*PBS= Pharmaceutical Benefits Scheme, MBS= Medicare Benefits Schedule, AR-DRG= Australian-refined Diagnostic Related Groups, ICU= Intensive Care Unit*

##### Supplementary Table 16: Cost per patient category

| Patient Category | Total Cost (2019 $AU) | Total Cost (2019 $AU) |
|---|---|---|
| Died | 24,665 | [deaths not costed separately in final PMSLT; rather deaths assumed to have cost of category below from whence they arose] |
| Treated in ICU | 64,238 | $44,641 |
| Non-ICU hospitalisation | 20,613 | $14,324 |
| Symptomatic case, no hospitalisation | 83 | $57.68 |
| Infected case with no symptoms | 0 | 0 |

ICU= Intensive care unit

#### Appendix 4 Road Traffic Crash

COVID-19 restrictions may result in lower road traffic deaths by reducing the number of people leaving their homes. Simultaneously, such restrictions have been linked to higher car and bike usage on roads, compared to public transport use as individuals aim to socially distance. Similarly, lower road congestion may be linked to higher speeds and potentially higher road traffic deaths. To estimate the effect of COVID-19 restrictions on road traffic deaths in Victoria, we obtained COVID-19 mobility data for January to October 2020 from Apple, which describes the activity of users seeking driving routing directions. This data was supplemented by the data on the road traffic deaths on Victorian roads from January through September 2020, from the Australian Bureau of Infrastructure and Transport Research Economics.

**Supplementary Figure 4: Apple mobility index for Victoria: Driver routing requests (100 = January Baseline)**

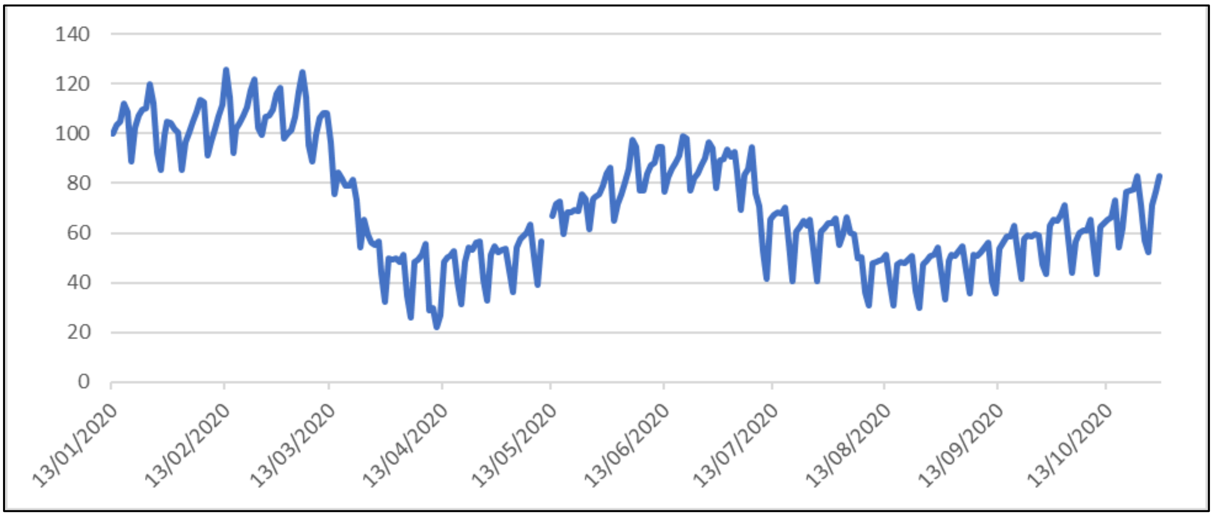

**Supplementary Figure 5: Victorian Traffic Fatalities by Month (Jan-Sep)**

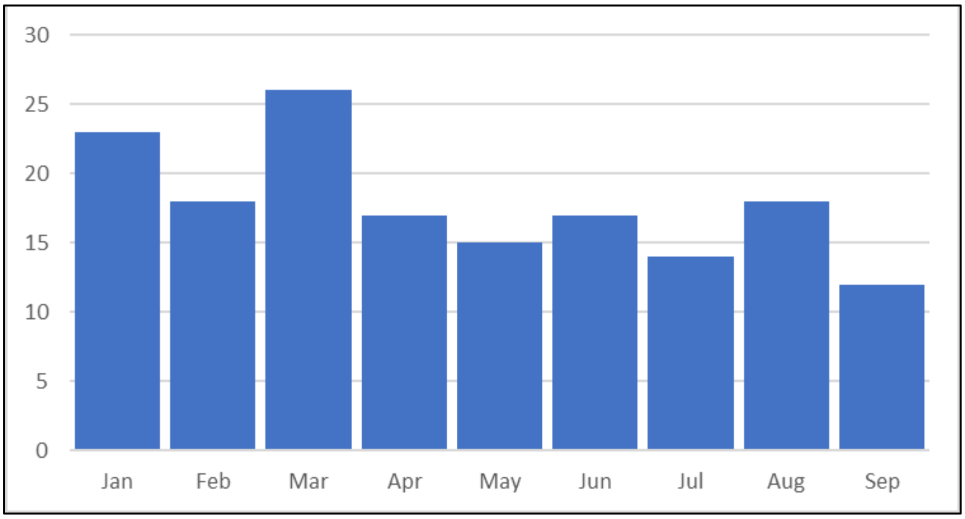

The data was used to calculate the association of a change in the driving mobility index on road traffic deaths using a Poisson regression. A 1-point increase in the mobility index causes a 0.45% increase in road traffic deaths, although the result is not statistically significant. The results of the regression are presented in Supplementary Table 17.

##### Supplementary Table 17: Association of Apple Driving Mobility Indices on Road Traffic Deaths

| | (1)<br>Number of Fatalities |
|---|---|
| Mobility Index | 0.00453<br>(1.28) |
| Day of Week Dummies | YES |
| Observations | 63 |

t statistics in parentheses

Next, the average Apple driving mobility index was calculated for each stage in VIC through calculating averages using data on restrictions available in Supplementary Table 18. The data was used to calculate the resulting change in road traffic deaths.

##### Supplementary Table 18: Apple mobility driving index for stages 2, 3 and 4 of COVID-19 policy restrictions

| **Stage** | **Time** | **Average Apple Mobility Index (100 = Jan 13 Baseline)** | **Change in RTC from Baseline** |
|---|---|---|---|
| 4 | 2nd August – 14th September | 47.68 | -23.6% (+12.6% to -59.9%) |
| 3 | 30th Mar – 13th May; 6th July – 2nd August | 51.57 | -21.93% (+11.3% to -55.5%) |
| 2 | 13th May – 27th June | 80.7 | -8.74% (-22.13% to 4.6%) |

Stages 1 and 1b are counterfactual. To approximate these mobility indices, we scaled the mobility indices to 0.488 the percent change in GDP (9.42% GDP loss in stage 2 = 19.3% GDP change) from Australian and Victorian Treasuries (See Supplementary Table 7).

##### Supplementary Table 19: Estimated changes in RTC rates for stages 1b and 1a

| Stage | GDP Change | Mobility Index | RTC |
|---|---|---|---|
| 1b | .535b/week = 6.95% | 96.6 | -1.53% (-3.8% to 0.8%) |
| 1a | .5bn/week = 6.5% | 96.8 | -1.45% (-3.6% to 0.77%) |

A limitation of the methodology based on using observational data is that the initial response to Stage 3 restrictions was stronger than later in July, this may be due to the lower national cases of COVID-19 when the second wave. The effect of Stage 3 on RTC cannot be disentangled from this behavior response to the initial wave and may therefore be overestimated.
